# Patient-Expressed Needs, Burdens and Barriers in Taiwanese Online Health Discussions: A Taiwan-Contextualized Friction Framework Applied With Large Language Model–Based Social Listening

**DOI:** 10.64898/2026.11.17.26365957

**Authors:** Zhi-Yong Liu, Tsao Chieh, Chang-Fu Kuo

**Affiliations:** Center for Artificial Intelligence in Medicine, Chang Gung Memorial Hospital, Taoyuan, Taiwan; Department of Internal Medicine, New Taipei Municipal TuCheng Hospital, New Taipei City, Taiwan; Division of Rheumatology, Allergy and Immunology, Chang Gung Memorial Hospital, Taoyuan, Taiwan; School of Medicine, Chung Shan Medical University, Taichung, Taiwan; School of Medicine, Chang Gung University, Taoyuan, Taiwan

**Keywords:** social media, patient experience data, patient-focused drug development, unmet needs, caregivers, natural language processing, large language models, content analysis, Taiwan, national health insurance

## Abstract

**Background:** Patients’ difficulties with illness and care (not understanding a result, distress, daily restrictions, not knowing where to go, family strain) may not surface in clinical encounters or records. Regulatory guidance on patient-focused drug development recognizes social-media listening as a source of patient experience data, but no systematic framework exists for Traditional Chinese discussions in Taiwan, and keyword pipelines measure such content poorly.

**Objective:** To characterize patient-expressed needs, burdens and barriers (“frictions”) in Taiwanese online health discussions with an eight-domain, Taiwan-contextualized framework, identify whose frictions they are, and assess how securely each is measured.

**Methods:** From a 13.6-million-unit archive of the PTT forum and three ask-a-physician services, seeded samples of 9,131 pipeline-flagged units and 1,000 unflagged forum articles (10,130 distinct units) were read in full by a large language model under rubrics for relevance, author (patient or family member) and eight domains: information gap, emotional burden, lifestyle burden, medication experience, system navigation, access barrier, caregiver burden and cost burden. Profiles were reported separately for forum articles and physician submissions and checked against a second vendor, two physicians’ independent labels of 160 accepted and rejected units, and the archive’s keyword pipeline.

**Results:** Among 2,395 model-confirmed forum articles, 83.1% voiced at least one friction: information gap 59.5% (physicians 86%–93%), emotional burden 39.0%, medication experience 24.1%, lifestyle burden 22.2% and system navigation 16.3%; access barrier, caregiver burden and cost burden were each below 7%. Of 3,105 submissions, nearly all were questions and 53.8% also carried a burden. Navigation rarely concerned insurance (9.1%). Family-authored forum articles (14.5%) more often voiced emotional burden (54.9% vs 36.3%), navigation (34.8% vs 13.2%; instrument-dependent) and access barriers (16.4% vs 4.7%), and patient-authored articles lifestyle burden (24.8% vs 6.9%); caregiver burden came with emotional burden in 86.0% of articles voicing it. Emotional and lifestyle burden were commoner in forum articles than submissions under the model, a second vendor and both physicians; medication experience and navigation were instrument-dependent, and physicians disagreed on cost (32% vs 4%). The keyword pipeline had stored its information-gap output for 29.9% of model-confirmed reference-stratum units and over-counted insurance navigation 2.9-fold; its relevance flag was 50.5% precise for forum articles and reached an estimated 42.9% of patient-voice articles.

**Conclusions:** Systematic listening organized Taiwanese patients’ and families’ online accounts into eight clinically interpretable friction domains, led by the need to understand and by emotional burden, with family-authored posts more often voicing navigation and access difficulty. These expressed concerns may not surface in routine encounters; they identify candidate targets for patient-centred communication and services, which require evaluation. Some domain levels depended on the reader, and keyword pipelines missed or distorted them, so such work needs full-text reading, explicit definitions and clinician checks.

## Introduction

Living with illness creates difficulties that patients do not always raise in a consultation or that a structured record does not capture: not understanding a result or what to do next, distress, restrictions of diet, work or sleep, uncertainty about which service to approach, costs, and strain on the family members who help. Patients and families describe these difficulties in public online channels, and patient-voice analysis now informs pharmacovigilance, patient-focused drug development and health policy [1–4]. Regulators recognize the passive observation of social media as a source of patient experience data, and best-practice guidance asks that whole forums be collected within platform terms, users pseudonymized, patients identified before coding, concepts coded against explicit definitions, automated coding validated against human annotation, and filtering steps reported [5–7]. We use *friction* for these recurring difficulties with illness and with the health system. A framework that names the frictions, measures how often they are voiced and by whom, and states how securely each is measured is a basis for patient-centred communication and services.

What patients struggle with is usually studied one disease, platform and construct at a time, and Taiwanese and Chinese-language listening studies have been single-community, small-scale or unvalidated [8–15]. Taiwan’s setting shapes how frictions are expressed: a single-payer National Health Insurance (NHI) system whose vocabulary (NHI, reimbursement, self-pay) saturates health talk [16,17]; direct access to hospitals and specialists, which turns choosing where to go into a task of its own; and family members who often write on a patient’s behalf. Patients speak on PTT, a non-commercial, academically operated bulletin board founded in 1995 and an established research source [18], and in ask-a-physician services, including the Ministry of Health and Welfare (MOHW) Taiwan e-Hospital. We found no systematic framework of patient frictions for Traditional Chinese.

Such a framework needs measurement that can be trusted. Production pipelines lean on keyword classifiers that over-count passing mentions [19,20] and under-count paraphrase [21], and the relevance flag that defines who counts as a patient can both include wrong units and miss real ones [22,23]. Large language models (LLMs) can read full text and match crowd annotators on many tasks [24–27], although they trail fine-tuned classifiers on some health benchmarks [28] and require validation against human judgment and stability reporting [29–31]. What a post contains also depends on where it is written: expert-facing services concentrate informational and treatment-seeking content, whereas peer spaces elicit experience-sharing and emotional support [32–34], and patients attach history, medication details and emotion to consultation questions [8,9,35]. A friction profile must therefore be reported by source rather than pooled.

Following the collection, selection and coding workflow of the patient-listening guideline [5,6], we applied an eight-domain friction framework, contextualized for Taiwan, to the full text of a sample of Taiwan’s online patient voice. We asked which frictions patients and families express, whose frictions they are (the patient’s or a family member’s), which frictions occur together, and how securely each domain can be measured by an LLM, a second vendor and two physicians. Secondarily, we asked how the source (a general-purpose forum or a physician service), the pipeline’s relevance flag and its keyword classifier change the profile.

## Methods

### Study Design and Data Source

This observational, cross-sectional content analysis is reported according to STROBE, with TRIPOD-LLM items for the LLM instrument [36,37] (Multimedia Appendix 3), and is organized along the collection, selection and coding workflow of the patient-listening guideline [6]. Posts were collected from Taiwanese public platforms by rate-limited crawlers between 2024 and 2026, covering material published from 1995 onward. Collection was exhaustive at the board or site level, not keyword filtered; disease and construct tags were applied after collection. Author identifiers were irreversibly hashed (salted SHA-256) at collection; publication dates, boards or departments and hashed author identifiers were joined to every analysed unit.

### Platform Governance

We audited each platform’s live terms of service, robots.txt directives and licences (verbatim snapshots archived 16 August 2026) before any corpus statistic was computed. Four platforms whose terms did not prohibit non-commercial academic analysis were included: PTT (ptt.cc; its user-content licence bars republication, so no content is reproduced), KingNet (kingnet.com.tw; robot-compliant collection), Taiwan e-Hospital (MOHW open-government data declaration) and tcm.tw (Chinese-medicine questions and answers). Dcard, BabyHome and 5914, whose terms or robots.txt prohibit automated collection, and unverifiable patient-association sites were excluded (Table S1 in Multimedia Appendix 1).

### The Friction Framework

The eight domains were defined by the investigators before this study, from the concepts of patient experience, burden of living with a condition, treatment experience and unmet need in patient-focused drug development guidance and the patient-listening guideline [5–7], and from clinical experience; they were not derived inductively from these posts. Each is a binary label, assigned only when the friction is present for the author and stated in the text (passing mentions do not count): information gap (seeks information or advice, asks a clinical question, or expresses diagnostic or treatment uncertainty); emotional burden (illness-tied distress); lifestyle burden (illness-driven restriction of diet, activity, work or sleep); medication experience (first-hand experience, efficacy doubt or side effects); system navigation (working through insurance coverage, eligibility or paperwork, or deciding which hospital, specialty or physician to approach); access barrier (concrete difficulty reaching care, such as appointments, referral, distance or waiting); caregiver burden (the author helps a patient and voices that burden); and cost burden (actual financial strain from care costs). The framework was contextualized for Taiwan in three respects: rubrics were applied to Traditional Chinese text, and insurance vocabulary counted as navigation or cost only when the author worked through it or was strained by it; navigation included choosing a hospital, specialty or physician, a frequent task where patients approach specialists directly; and the author was coded separately from the person whose health was discussed, so that a family member’s own strain (caregiver burden) was distinguished from the patient’s frictions. A post may carry several domains; domains are reported separately and are not summed into a severity score. The verbatim rubrics are in Multimedia Appendix 2. The framework’s coverage of difficulties outside the eight domains was not audited in this study (see Limitations).

### Units of Analysis and Sources

On PTT an article is an authored post, and a push is a one-line comment appended beneath an article by another user that the pipeline indexes and flags as a separate unit; pushes are reported as their own unit type and the open-forum source is PTT articles. On KingNet, Taiwan e-Hospital and tcm.tw every unit is a submission to a physician, a question by design. The three services share that property (99.0–99.9% questions) and are pooled with design weights for their flagged populations (largest weighted-minus-unweighted difference 0.02 points; per-site rates in Table S2 in Multimedia Appendix 1). Friction profiles are reported separately for forum articles and physician submissions; no rate pooled across them is reported as an estimate. KingNet units carry the physician’s reply; a question-only re-read was a sensitivity analysis.

### Study Population and Sample

The archive comprised 13,568,496 distinct units: 12,628,592 from PTT, 348,058 from KingNet, 309,840 from tcm.tw and 282,006 from Taiwan e-Hospital. Of these, 1,029,728 were flagged as patient voice by the pipeline, and 939,007 flagged units with a body of at least 20 characters formed the sampling frame. The sample had three strata, all drawn with fixed seeds (Table 1): a random sample of the flagged frame (3,631 units after the terms audit; the reference stratum), which also carries the single-unit reference read described below; random oversamples of flagged forum articles (4,000, from the 190,763 flagged articles on the 41 boards represented in the sample) and of flagged submissions (1,500, in proportion to each service’s flagged population), to give the source comparison precision; and 1,000 unflagged articles from the same boards (525,295 in the archive) to estimate the flag’s recall (Figure 1). Because the oversample was restricted to the boards represented in the random stratum, source contrasts were repeated on the random stratum alone. The oversamples were drawn from the frame after the random stratum had been removed. In all, 9,131 flagged units were read (4,738 articles, 1,187 push comments and 3,206 submissions); with the unflagged articles, 10,130 distinct units were annotated (one forum article appeared twice in the random stratum as two frame entries of the same post; both entries are counted in Table 1, which changes no reported rate materially). The other 13.6 million archive units were not read. Throughout, “confirmed” means confirmed as patient voice by the model’s relevance read (model-confirmed), not by a human.

**Figure 1.**
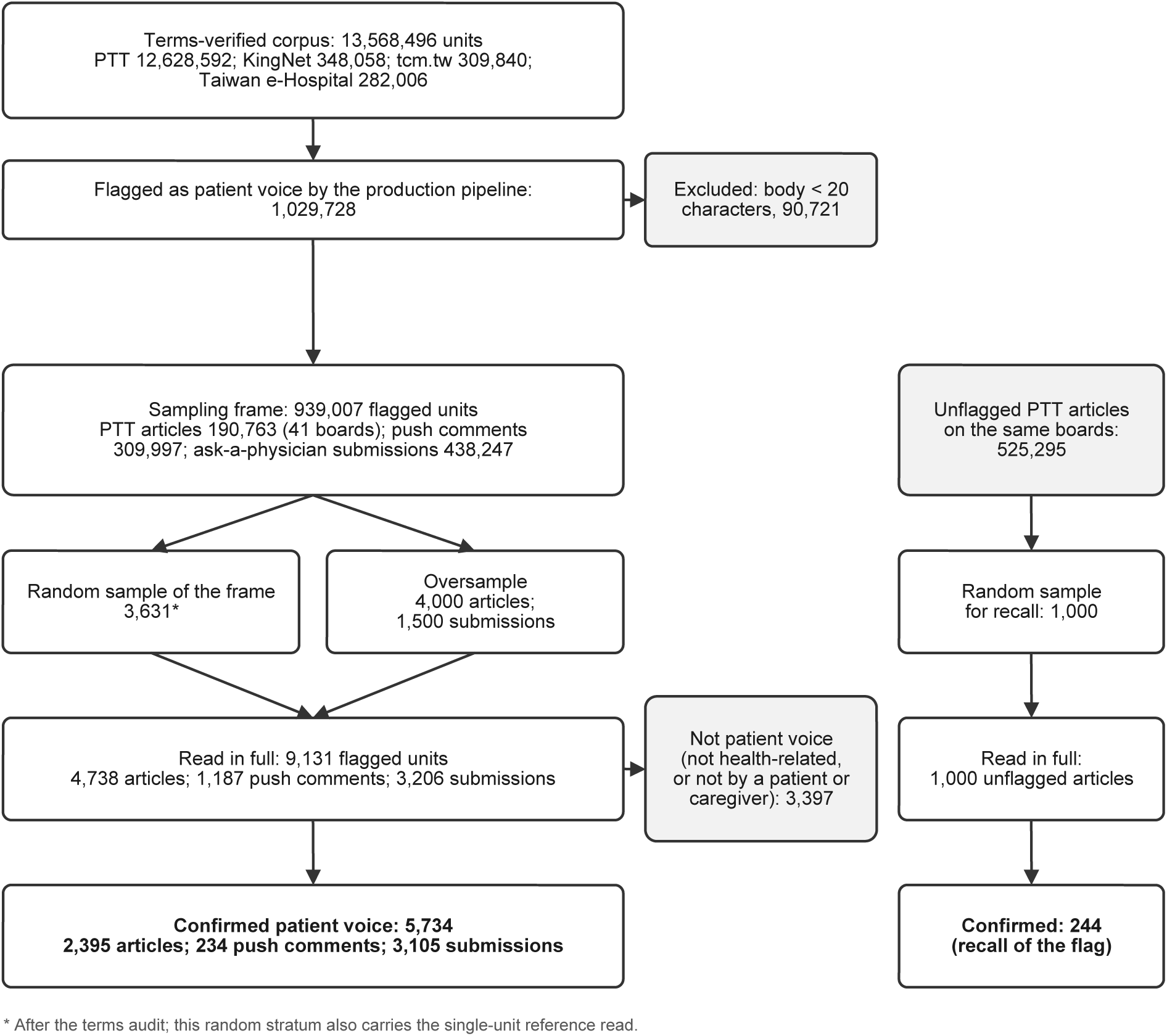
Flow of units from the corpus to the analysed sample. Counts at each step: the terms-verified corpus, units flagged as patient voice by the production pipeline, the sampling frame of flagged units with at least 20 characters, the three seeded strata, units read in full, and units confirmed as patient voice by the relevance read; the unflagged articles read to estimate the flag’s recall are shown on the right.

**Table 1.** The terms-verified corpus, the sample, and precision and recall of the production patient-voice flag.

| **Platform and unit type** | **Source type** | **Units in corpus** | **Frame: flagged units ≥20 characters (last row: unflagged articles)** | **Read** | **Confirmed** | **Precision of the flag, % (95% CI)** | **Estimated confirmed patient voice, thousands (95% CI)** |
|---|---|---|---|---|---|---|---|
| PTT articles | Open forum | 12,628,592 (all PTT units) | 190,763 | 4,738 | 2,395 | 50.5 (49.1–52.0) | 96.4 (93.7–99.1) |
| PTT push comments | Open forum (comments) | — | 309,997 | 1,187 | 234 | 19.7 (17.5–22.1) | 61.1 (54.4–68.4) |
| KingNet | Ask-a-physician | 348,058 | 187,497 | 1,371 | 1,318 | 96.1 (95.0–97.0) | 180.2 (178.1–181.9) |
| Taiwan e-Hospital | Ask-a-physician (government) | 282,006 | 180,652 | 1,317 | 1,293 | 98.2 (97.3–98.8) | 177.4 (175.8–178.4) |
| tcm.tw | Ask-a-physician (Chinese medicine) | 309,840 | 70,098 | 518 | 494 | 95.4 (93.2–96.9) | 66.9 (65.3–67.9) |
| Total, flagged |  | 13,568,496 | 939,007 | 9,131 | 5,734 | — | 582 (574–590) |
| PTT articles not flagged | Open forum | — | 525,295 | 1,000 | 244 | Recall of the flag on articles: 42.9 (40.2–45.7) | All patient-voice articles: 225 (210–239) |

Flagged = identified as patient- or caregiver-authored by the production pipeline; the sampling frame is flagged units with a body of at least 20 characters (90,721 shorter flagged units, almost all push comments, were excluded). PTT article frames are the boards represented in the sample. Precision = share of flagged units read that the relevance read confirmed as health-related and patient- or caregiver-voiced (Wilson 95% CI); estimated model-confirmed counts apply each unit type’s precision interval to its flagged frame; the total’s interval combines the unit-type strata as independent binomial estimates. Recall = confirmed flagged articles ÷ all confirmed articles in the archive, from the flagged and unflagged frames (delta-method CI on the log-odds scale; Monte Carlo 40.3–45.8). Platforms that failed the terms audit (Dcard, BabyHome, 5914) and unverifiable platforms were excluded before analysis; the terms basis of each included platform is in Table S1 in Multimedia Appendix 1.

### Patient-Voice Confirmation and Author

Every sampled unit was re-read in full under a fixed relevance rubric: does it concern a specific person’s health, and whose voice is speaking (patient; caregiver, that is, a family member or other person writing about or for the patient; clinician; media; other). A unit was confirmed patient voice if it was health-related and written by a patient or a family member; we call the latter family-authored. Flag precision is the confirmed share of flagged units, by unit type; the flag’s recall on articles is the confirmed flagged share of all confirmed articles in the archive, estimated as N_F_p_F_ ÷ (N_F_p_F_ + N_U_p_U_) from the flagged (F) and unflagged (U) article frames, with a delta-method interval on the log-odds scale and a Monte Carlo check. The confirmed subset is the denominator for every rate; rates on all flagged units are a sensitivity analysis.

### Full-Text Friction Measurement

Each unit (title and body, truncated at 6,000 characters) was read under the friction rubric by gpt-6.1-sol (OpenAI; Codex command-line interface; 23 October 2026) in documents of 25 units that carried both verbatim rubrics; the model returned one JSON (JavaScript Object Notation) object per unit with its identifier echoed, documents were re-read until complete, sampling settings were provider defaults with no seed, and units were placed in documents as seeded mixtures of unit types and years. Only 25 articles (0.5%), and no other unit, exceeded 6,000 characters. Model and document size were chosen in a pilot of four models at 25, 100 and 300 units per document; the pilot served only these choices and is not reported further. On the services every submission is a question by design, so the informative measure is the burden that accompanies the question. Prompts, except that of the strict NHI-navigation read, are in Multimedia Appendix 2.

### Agreement and Replication

The instrument was compared with three references (a single-unit reference read, earlier physician reference sets and a second-vendor replica), and separately with two physicians (see Physician Comparison). First, the 3,631-unit stratum had been read unit by unit by GPT-5.5 (OpenAI; OpenRouter route openai/gpt-5.5; June–August 2026; single user message; provider-default temperature); agreement with this single-unit reference read measures consistency across reading formats of the same vendor, not accuracy against human judgment. Second, physician labels exist for information gap on a 50-unit Taiwanese reference set (two physicians; κ 0.79 between them on 29 units; drawn from a 91-post physician pool that included 19 posts from platforms later excluded or unrecorded) and for NHI navigation on a separate set of 92 units. Third, every unit was read end to end by a second vendor (Claude Sonnet 5.5, Anthropic) with the same documents and rubrics. A domain was pre-specified as instrument-dependent if its replica κ among units confirmed by both instruments was below 0.60, or if its services-minus-articles contrast changed sign or significance under the replica; such domains are reported descriptively.

### Physician Comparison

Two physician authors (CT and CFK; CFK also designed the instrument) independently rated 160 units in a bilingual worksheet that reproduced the instrument’s definitions of relevance, voice and the eight domains; the worksheet contained no model output, units were presented in shuffled order, and neither physician saw the other’s ratings. Units were drawn with a fixed seed from the full-text annotation (all units read in full: the flagged sample and the unflagged articles): 95 instrument-confirmed units at random within unit type (the random sample: 37 articles, 17 push comments and 41 submissions), 30 units the instrument had not confirmed (15, 10 and 5), and 25 units enriched for rare domains and 10 for disagreement between the instrument and the replica. Each physician labelled relevance and voice for every unit and the domains for units they judged patient or caregiver voice. Each physician personally read every unit and entered the labels; no labels were pre-filled by a model or other software. There was no adjudication. Both physicians were aware of the study aims and one designed the instrument, so this comparison is a check against non-independent raters, not validation against a reference standard. The instrument was compared with each physician separately and benchmarked against agreement between the physicians, using Cohen κ and Gwet’s AC1, which is robust to skewed prevalence [38]; κ intervals were not computed where either source had fewer than five positives. Weighting sampled units to their unit type’s count in the full-text annotation, we estimated, by unit type, the share of instrument-confirmed units each physician confirmed, the share of unconfirmed units each physician confirmed, the resulting share of physician-identified patient voice that the instrument captured, and domain rates and services-minus-articles differences under each physician’s labels, with percentile intervals from 2,000 resamples within strata. Because the full-text annotation combines a flagged sample with unflagged articles, these estimates describe the units read, by unit type, not the archive.

### Keyword Classifier Benchmark

The archive’s production pipeline relies on a rule-based heuristic (version 2.4) using Traditional Chinese keyword sets with jieba segmentation that assigns relevance classes and, for patient-voice units, friction aspects including an information gap. Its measurement was benchmarked on the reference stratum, where its outputs are stored, on two domains chosen a priori to span the breadth spectrum: information gap (broad), against the full-text model label, and NHI navigation (narrow; genuine engagement with the insurance system, distinguished from passing mentions), against a strict single-unit model read. Both references are model labels, not adjudicated human truth. The pipeline’s stored information-gap output was missing for many units, apparently those outside its aspect-processing scope (on PTT, mostly older posts); we report the share with a stored output and compare rates both as stored (missing treated as absent, as in the production outputs) and on units with a stored output.

The keyword instrument for NHI navigation is a 24-term NHI lexicon (Multimedia Appendix 2); where the pipeline’s detection and measurement lexicons overlap, measurement is circular: all but one of 2,930 aspect-labelled units containing the term for self-pay were labelled cost-positive by the pipeline, and all 908 containing the term for referral system-navigation-positive, so such pipeline labels were not used.

### Statistical Analysis

Rates carry Wilson 95% CIs [39]; differences carry Newcombe 95% CIs [40] and rate ratios log-method CIs. The friction profile is reported by source (forum articles; submissions), with push comments as a separate unit type. In the source comparison, the seven burden contrasts and the any-burden contrast were tested with Fisher’s exact test and Holm correction across the eight contrasts in each table [41], and only differences surviving correction are called significant. Heterogeneity across services used χ² tests. Dependence of pooled rates on source composition is shown by the pooled rate implied by each article share and by counting domains whose pooled value at the projected model-confirmed article share in the flagged frame (18.5%) lies outside both source CIs. Agreement is Cohen κ with percentage agreement, positive and negative specific agreement and stated denominators [42–44], with bootstrap CIs resampling documents (batched reads) or units (physician sets). Sensitivity analyses of the source contrast: the random stratum alone, the board-restricted oversample alone, Mantel–Haenszel differences across common length tertiles, exclusion of appearance and fitness boards, a topic-matched psychiatry comparison, two-of-three consensus labels on the reference stratum, the second-vendor replica and units published in 2020 or later; also author-clustered intervals and services without tcm.tw or KingNet. The domain summary across readers, the comparison of patient-authored with family-authored posts and the co-occurrence of four domain pairs (pairs fixed before computing) were specified after the primary results had been inspected; they are descriptive and carry no multiplicity adjustment. All numbers were generated by scripts from per-unit labels and re-derived by a separate verification script.

### Ethical Considerations

The Institutional Review Board of Chang Gung Memorial Hospital approved the study (No. 202601393B0) and waived the requirement for informed consent because the posts are public and pseudonymous. We analysed lawfully public, non-interactive information under Taiwan’s Personal Data Protection Act Articles 6, 19 and 20 and the fair-use provision of the Copyright Act; identifiers were pseudonymized at collection and no user content is reproduced. Following Eysenbach and Till [45], all four platforms are readable without registration, none is a closed support group and the research was passive; because submissions to physicians may carry a higher expectation of privacy, reporting is aggregate only. Post text, with author identifiers and any names, contact details or other identifiers within post bodies removed, was sent solely for labelling to OpenAI (through the Codex command-line interface, and through the OpenRouter application programming interface for the earlier single-unit reference reads) and to Anthropic, under provider and intermediary account settings that exclude customer content from model training; the authors did not use post text to train or fine-tune any model. Of the 91 posts in the Taiwanese physician reference pool, 19 came from platforms later excluded by the terms audit or had no recorded platform; they were labelled before the audit concluded and contribute to agreement statistics only. No participant was contacted and no compensation was provided [46].

## Results

### Confirmed Units

Of 9,131 flagged units read, 5,734 were confirmed as health-related and patient- or family-authored: 2,395 forum articles, 234 push comments and 3,105 submissions (KingNet 1,318, Taiwan e-Hospital 1,293, tcm.tw 494). Confirmed articles came from 2,274 authors on 33 of the 41 boards in the frame (median length 482 characters; median year 2015), and submissions from 73 departments (median length 170; median year 2012; Table S3 in Multimedia Appendix 1). Family members wrote 14.5% of confirmed articles (348) and 14.0% of submissions (436).

### The Friction Profile

Among confirmed forum articles, 83.1% voiced at least one friction and 63.4% a burden beyond a question (Table 2; Figure 2). The need to understand led: 59.5% sought information, asked a clinical question or expressed uncertainty about a diagnosis or treatment. Emotional burden followed (39.0%), then medication experience (24.1%), lifestyle burden (22.2%) and system navigation (16.3%). Access barriers (6.4%), caregiver burden (6.6%) and cost burden (2.9%) were each voiced in fewer than 7% of articles. Among submissions, 99.6% were questions by construction, and 53.8% carried a further burden with the question: medication experience 25.0%, emotional burden 20.4%, system navigation 15.0% and lifestyle burden 11.9%; access, caregiver and cost burden were each 2.5% or less. Push comments, one-line reactions, carried little: 23.9% voiced any friction, mostly medication remarks (12.0%), and their confirmation was instrument-dependent (replica κ 0.48; Table S5).

**Figure 2.**
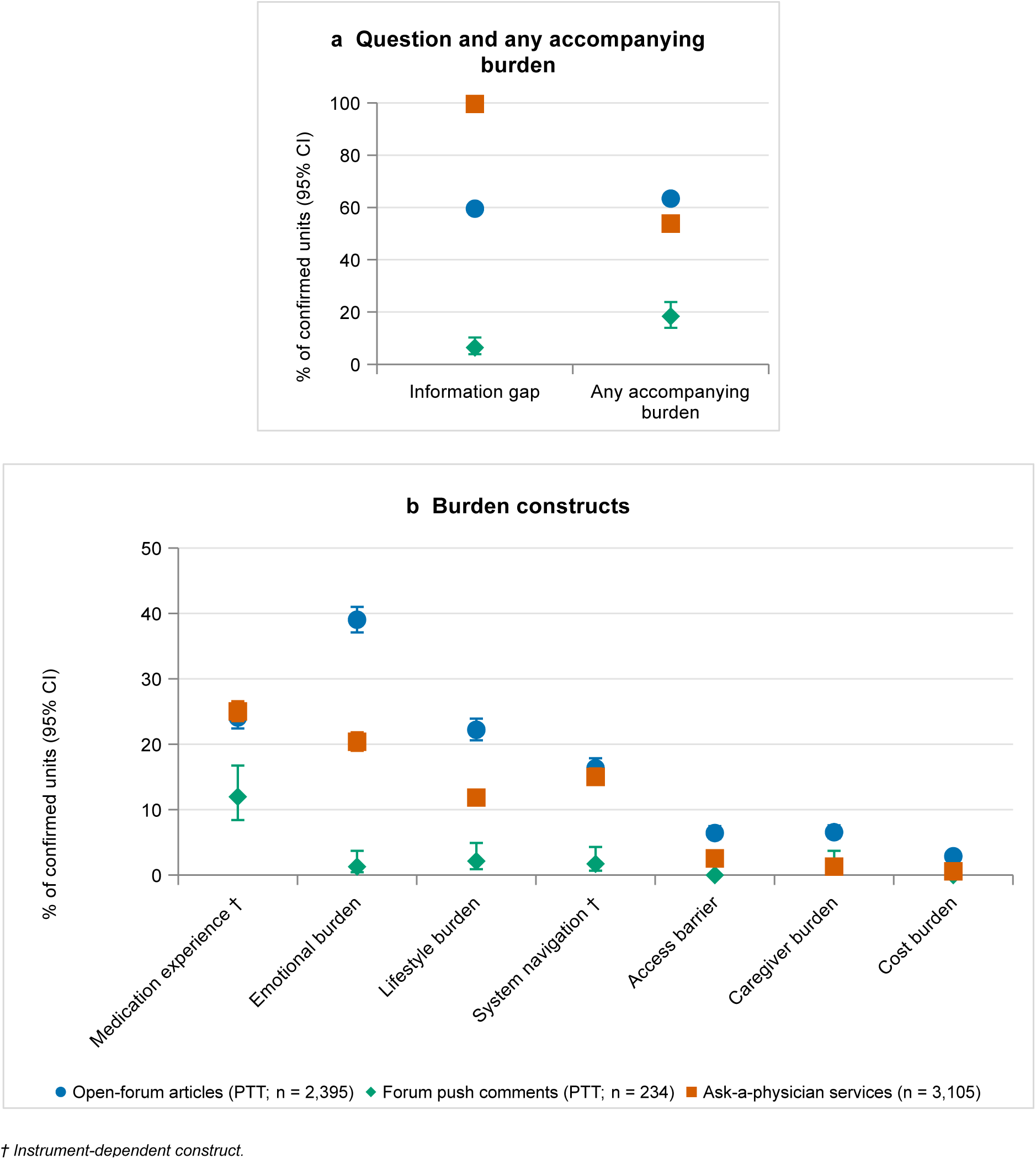
Friction map by unit type. Rates with Wilson 95% CIs of the eight constructs and any burden among confirmed forum articles (PTT; n = 2,395), forum push comments (n = 234) and ask-a-physician submissions (n = 3,105); the information-gap construct is satisfied by format on the services. † Instrument-dependent construct.

**Table 2.**
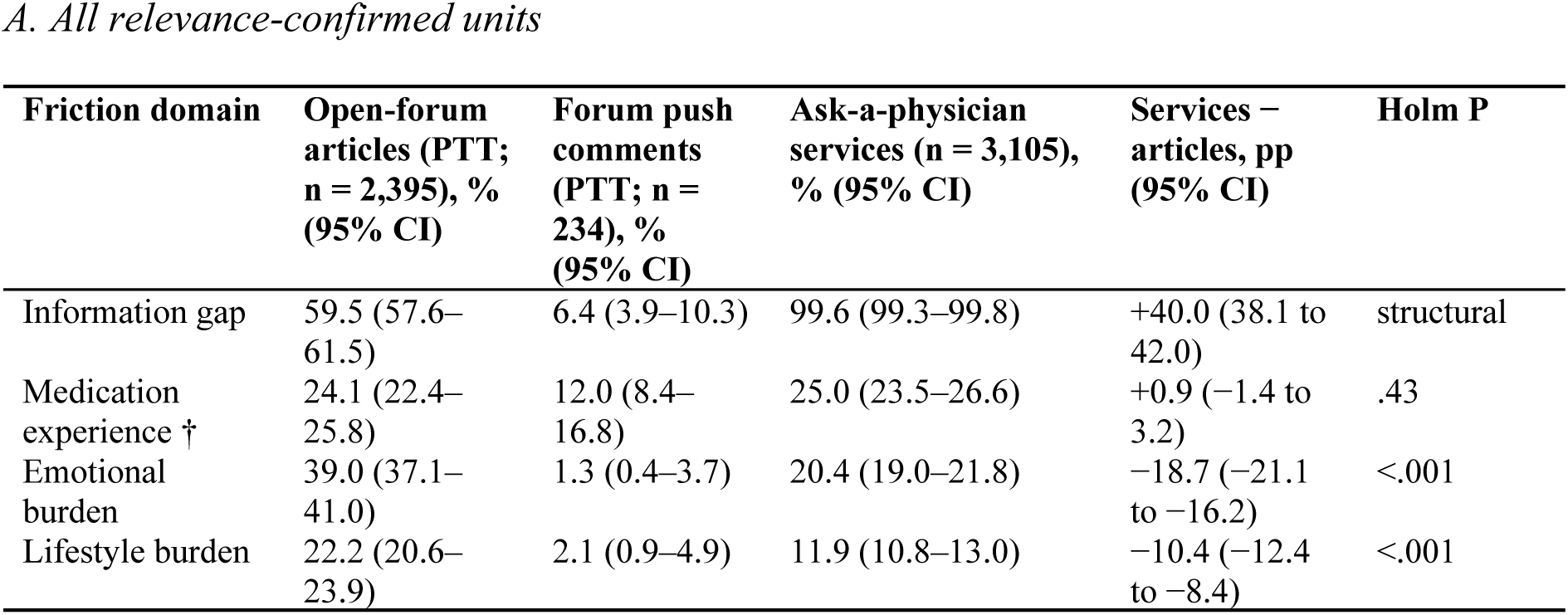
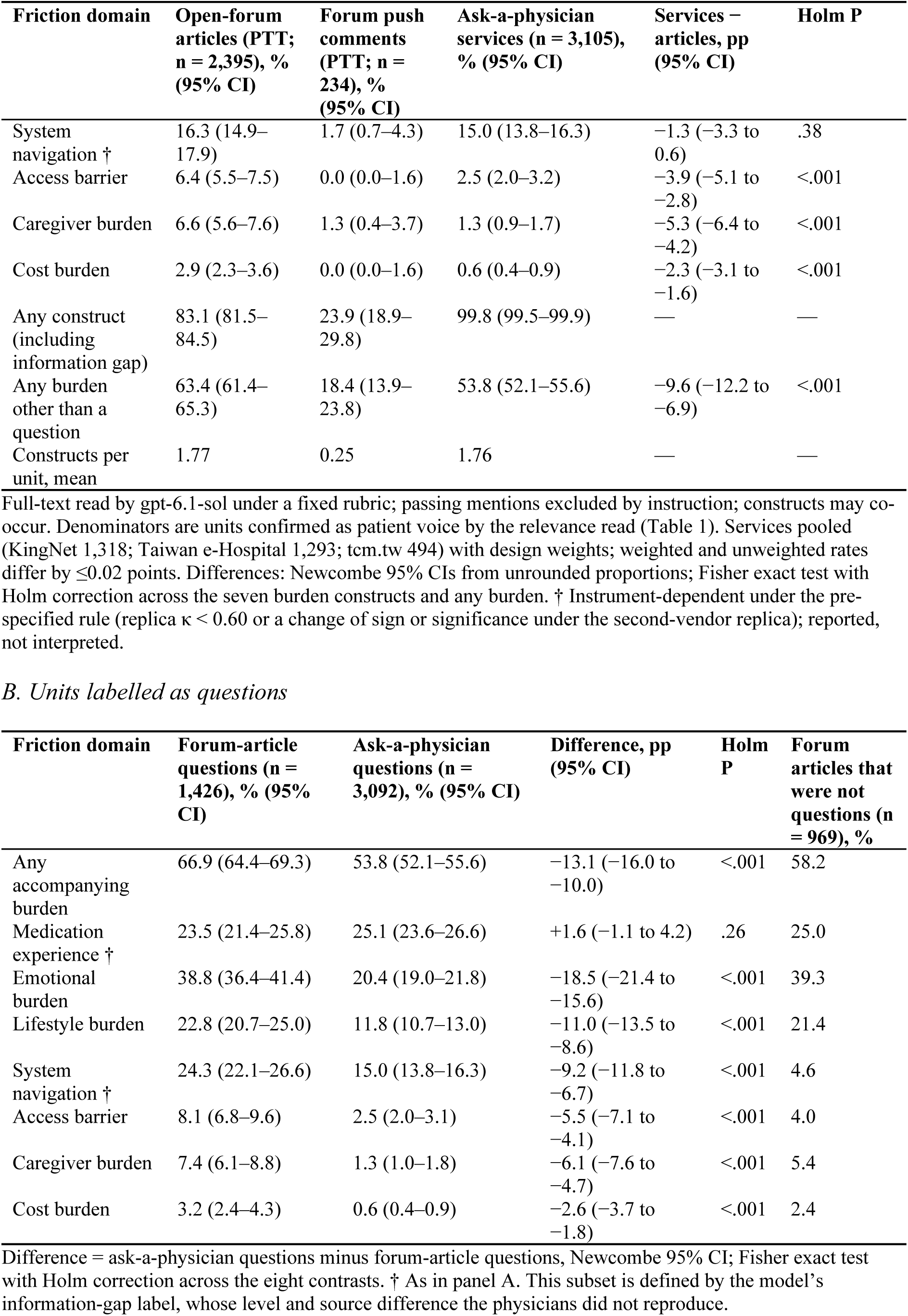
The friction profile by unit type, and the burden accompanying a question.

| **Friction domain** | **Open-forum articles (PTT; n = 2,395), % (95% CI)** | **Forum push comments (PTT; n = 234), % (95% CI)** | **Ask-a-physician services (n = 3,105), % (95% CI)** | **Services − articles, pp (95% CI)** | **Holm P** |
|---|---|---|---|---|---|
| Information gap | 59.5 (57.6–61.5) | 6.4 (3.9–10.3) | 99.6 (99.3–99.8) | +40.0 (38.1 to 42.0) | structural |
| Medication experience † | 24.1 (22.4–25.8) | 12.0 (8.4–16.8) | 25.0 (23.5–26.6) | +0.9 (−1.4 to 3.2) | .43 |
| Emotional burden | 39.0 (37.1–41.0) | 1.3 (0.4–3.7) | 20.4 (19.0–21.8) | −18.7 (−21.1 to −16.2) | <.001 |
| Lifestyle burden | 22.2 (20.6–23.9) | 2.1 (0.9–4.9) | 11.9 (10.8–13.0) | −10.4 (−12.4 to −8.4) | <.001 |
| **Friction domain** | **Open-forum articles (PTT; n = 2,395), % (95% CI)** | **Forum push comments (PTT; n = 234), % (95% CI)** | **Ask-a-physician services (n = 3,105), % (95% CI)** | **Services − articles, pp (95% CI)** | **Holm P** |
| System navigation † | 16.3 (14.9–17.9) | 1.7 (0.7–4.3) | 15.0 (13.8–16.3) | −1.3 (−3.3 to 0.6) | .38 |
| Access barrier | 6.4 (5.5–7.5) | 0.0 (0.0–1.6) | 2.5 (2.0–3.2) | −3.9 (−5.1 to −2.8) | <.001 |
| Caregiver burden | 6.6 (5.6–7.6) | 1.3 (0.4–3.7) | 1.3 (0.9–1.7) | −5.3 (−6.4 to −4.2) | <.001 |
| Cost burden | 2.9 (2.3–3.6) | 0.0 (0.0–1.6) | 0.6 (0.4–0.9) | −2.3 (−3.1 to −1.6) | <.001 |
| Any construct (including information gap) | 83.1 (81.5–84.5) | 23.9 (18.9–29.8) | 99.8 (99.5–99.9) | — | — |
| Any burden other than a question | 63.4 (61.4–65.3) | 18.4 (13.9–23.8) | 53.8 (52.1–55.6) | −9.6 (−12.2 to −6.9) | <.001 |
| Constructs per unit, mean | 1.77 | 0.25 | 1.76 | — | — |
| *B. Units labelled as questions* |  |  |  |  |  |
| **Friction domain** | **Forum-article questions (n = 1,426), % (95% CI)** | **Ask-a-physician questions (n = 3,092), % (95% CI)** | **Difference, pp (95% CI)** | **Holm P** | **Forum articles that were not questions (n = 969), %** |
| Any accompanying burden | 66.9 (64.4–69.3) | 53.8 (52.1–55.6) | −13.1 (−16.0 to −10.0) | <.001 | 58.2 |
| Medication experience † | 23.5 (21.4–25.8) | 25.1 (23.6–26.6) | +1.6 (−1.1 to 4.2) | .26 | 25.0 |
| Emotional burden | 38.8 (36.4–41.4) | 20.4 (19.0–21.8) | −18.5 (−21.4 to −15.6) | <.001 | 39.3 |
| Lifestyle burden | 22.8 (20.7–25.0) | 11.8 (10.7–13.0) | −11.0 (−13.5 to −8.6) | <.001 | 21.4 |
| System navigation † | 24.3 (22.1–26.6) | 15.0 (13.8–16.3) | −9.2 (−11.8 to −6.7) | <.001 | 4.6 |
| Access barrier | 8.1 (6.8–9.6) | 2.5 (2.0–3.1) | −5.5 (−7.1 to −4.1) | <.001 | 4.0 |
| Caregiver burden | 7.4 (6.1–8.8) | 1.3 (1.0–1.8) | −6.1 (−7.6 to −4.7) | <.001 | 5.4 |
| Cost burden | 3.2 (2.4–4.3) | 0.6 (0.4–0.9) | −2.6 (−3.7 to −1.8) | <.001 | 2.4 |

Full-text read by gpt-6.1-sol under a fixed rubric; passing mentions excluded by instruction; constructs may co-occur. Denominators are units confirmed as patient voice by the relevance read (Table 1). Services pooled (KingNet 1,318; Taiwan e-Hospital 1,293; tcm.tw 494) with design weights; weighted and unweighted rates differ by ≤0.02 points. Differences: Newcombe 95% CIs from unrounded proportions; Fisher exact test with Holm correction across the seven burden constructs and any burden. † Instrument-dependent under the pre-specified rule (replica κ < 0.60 or a change of sign or significance under the second-vendor replica); reported, not interpreted.
Difference = ask-a-physician questions minus forum-article questions, Newcombe 95% CI; Fisher exact test with Holm correction across the eight contrasts. † As in panel A. This subset is defined by the model's information-gap label, whose level and source difference the physicians did not reproduce.

Navigation rarely concerned insurance: on the reference stratum only 9.1% (95% CI 6.3–12.9) of navigation units engaged with the insurance system; under the domain’s definition the rest mainly concerned where to seek care (which hospital, specialty or physician), which was not coded separately. The model rarely labelled explicit financial strain, but the physicians disagreed widely on cost burden (one labelled it in 32% of the sampled forum articles they judged patient voice, the other in 4%; κ 0.21), so this low rate may reflect the instrument’s restrictive definition (actual strain, not price mentions) as much as what patients wrote; single-payer coverage, a younger forum population, self-pay cosmetic topics and reluctance to raise money with a physician are other explanations these data cannot separate. Because medication experience and navigation can be neutral (for example, choosing a hospital), a composite restricted to emotional, lifestyle, access, caregiver and cost burden, examined post hoc, was present in 51.4% (95% CI 49.4–53.4) of articles and 30.0% (28.4–31.7) of submissions.

The two sources carried different frictions. Compared with forum articles, submissions carried less emotional burden (−18.7 points), lifestyle burden (−10.4), caregiver burden (−5.3), access barriers (−3.9) and cost burden (−2.3), all Holm-significant and reproduced by the replica, and preserved in each sensitivity analysis of the full sources (Table S4 in Multimedia Appendix 1); medication experience and navigation did not differ. The broader any-burden difference (−9.6 points, 95% CI −12.2 to −6.9) was sensitive to analytic choices: −4.8 (95% CI −10.0 to 0.5) on the random stratum alone, −2.4 (95% CI −5.4 to 0.6) across length tertiles and −5.0 (95% CI −10.5 to 0.6) on consensus labels. Restricting both sources to questions, a secondary analysis defined by the model’s information-gap label, any accompanying burden was present in 66.9% of the 1,426 article questions and 53.8% of the 3,092 submission questions (Table 2B), so the frictions that accompany a question, rather than whether a question is asked, distinguish a forum article from a physician submission. Burden varied widely across boards: excluding the six appearance and fitness boards raised the article any-burden rate to 80.5% (Table S4), and psychiatry boards carried more emotional burden than psychiatry submissions (by 18.1 points; 95% CI 5.7 to 32.2). The services differed among themselves most on system navigation (range 13.3 points; Table S2 in Multimedia Appendix 1).

### Whose Frictions: Patients and Family Members

Family-authored posts voiced different frictions from patient-authored ones (Table 3A). On the forum, family-authored articles more often voiced emotional burden (54.9% vs 36.3%; difference +18.5 points, 95% CI +12.9 to +24.1), system navigation (34.8% vs 13.2%), access barriers (16.4% vs 4.7%) and an information gap (71.3% vs 57.5%), and 44.3% voiced the family member’s own caregiver burden. Patient-authored articles more often voiced lifestyle burden (24.8% vs 6.9%) and medication experience (25.6% vs 14.9%). In submissions, apart from caregiver burden, only the navigation (19.0% vs 14.3%), lifestyle (0.9% vs 13.6%) and medication (16.3% vs 26.5%) differences recurred, and family-authored submissions carried less burden overall (45.9% vs 55.1%; −9.2 points, 95% CI −14.2 to −4.2).

**Table 3.**
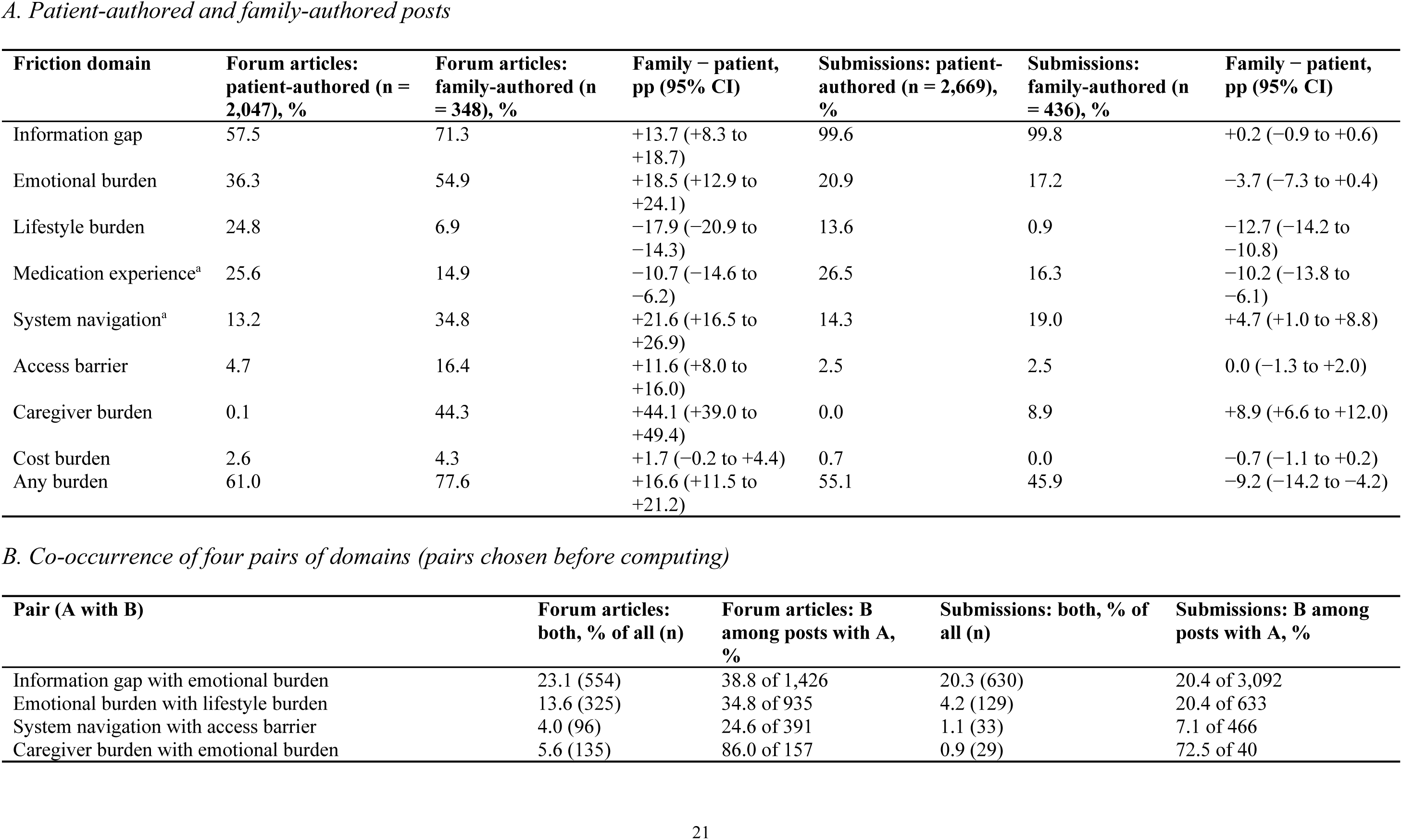
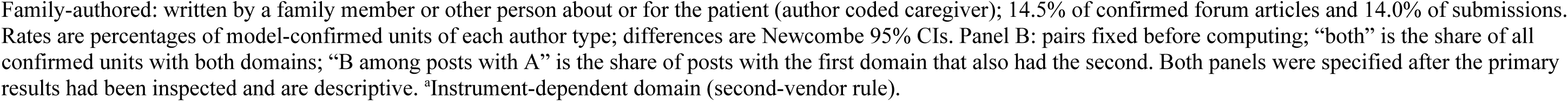
Whose frictions, and which occur together: patient- and family-authored posts, and co-occurrence of four domain pairs (model-confirmed units).

|  |  |  |  |  |  |  |
|---|---|---|---|---|---|---|
| *A. Patient-authored and family-authored posts* |  |  |  |  |  |  |
| **Friction domain** | **Forum articles: patient-authored (n = 2,047), %** | **Forum articles: family-authored (n = 348), %** | **Family − patient, pp (95% CI)** | **Submissions: patient-authored (n = 2,669), %** | **Submissions: family-authored (n = 436), %** | **Family − patient, pp (95% CI)** |
| Information gap | 57.5 | 71.3 | +13.7 (+8.3 to +18.7) | 99.6 | 99.8 | +0.2 (−0.9 to +0.6) |
| Emotional burden | 36.3 | 54.9 | +18.5 (+12.9 to +24.1) | 20.9 | 17.2 | −3.7 (−7.3 to +0.4) |
| Lifestyle burden | 24.8 | 6.9 | −17.9 (−20.9 to −14.3) | 13.6 | 0.9 | −12.7 (−14.2 to −10.8) |
| Medication experience<sup>a</sup> | 25.6 | 14.9 | −10.7 (−14.6 to −6.2) | 26.5 | 16.3 | −10.2 (−13.8 to −6.1) |
| System navigation<sup>a</sup> | 13.2 | 34.8 | +21.6 (+16.5 to +26.9) | 14.3 | 19.0 | +4.7 (+1.0 to +8.8) |
| Access barrier | 4.7 | 16.4 | +11.6 (+8.0 to +16.0) | 2.5 | 2.5 | 0.0 (−1.3 to +2.0) |
| Caregiver burden | 0.1 | 44.3 | +44.1 (+39.0 to +49.4) | 0.0 | 8.9 | +8.9 (+6.6 to +12.0) |
| Cost burden | 2.6 | 4.3 | +1.7 (−0.2 to +4.4) | 0.7 | 0.0 | −0.7 (−1.1 to +0.2) |
| Any burden | 61.0 | 77.6 | +16.6 (+11.5 to +21.2) | 55.1 | 45.9 | −9.2 (−14.2 to −4.2) |
| *B. Co-occurrence of four pairs of domains (pairs chosen before computing)* |  |  |  |  |  |  |
| **Pair (A with B)** | **Forum articles: both, % of all (n)** | **Forum articles: B among posts with A, %** | **Submissions: both, % of all (n)** | **Submissions: B among posts with A, %** |  |  |
| Information gap with emotional burden | 23.1 (554) | 38.8 of 1,426 | 20.3 (630) | 20.4 of 3,092 |  |  |
| Emotional burden with lifestyle burden | 13.6 (325) | 34.8 of 935 | 4.2 (129) | 20.4 of 633 |  |  |
| System navigation with access barrier | 4.0 (96) | 24.6 of 391 | 1.1 (33) | 7.1 of 466 |  |  |
| Caregiver burden with emotional burden | 5.6 (135) | 86.0 of 157 | 0.9 (29) | 72.5 of 40 |  |  |

Family-authored: written by a family member or other person about or for the patient (author coded caregiver); 14.5% of confirmed forum articles and 14.0% of submissions. Rates are percentages of model-confirmed units of each author type; differences are Newcombe 95% CIs. Panel B: pairs fixed before computing; “both” is the share of all confirmed units with both domains; “B among posts with A” is the share of posts with the first domain that also had the second. Both panels were specified after the primary results had been inspected and are descriptive. <sup>a</sup>Instrument-dependent domain (second-vendor rule).

### How Frictions Occur Together

Distress was as common with a question as without one: among forum articles with an information gap, 38.8% voiced emotional burden, against 39.3% of articles without one (Table 2B) and 39.0% of all articles (submissions 20.4% and 20.4%; Table 3B). Other frictions clustered. Among articles with emotional burden, 34.8% also voiced lifestyle burden (22.2% of all articles), and among those with system navigation, 24.6% also voiced an access barrier (6.4%; submissions 7.1% and 2.5%). Caregiver burden usually came with emotional burden (86.0% of articles and 72.5% of submissions with caregiver burden, against 39.0% and 20.4% overall).

### How Securely Each Domain Was Measured

Table 4 summarizes, for each domain, its level under the model, under the second vendor on the same units, and under each physician in the random sample, with agreement. On the 3,631 units read by both, the full-text instrument agreed with the single-unit reference read on confirmed status at κ = 0.89 (95% CI 0.88–0.91), on information gap at κ = 0.94 and on the other domains at κ = 0.66–0.89 (Table S5 in Multimedia Appendix 1); on Taiwan e-Hospital, where nearly every unit is confirmed, κ was 0.29 at 98.0% agreement, the prevalence paradox of κ [43,44]. The replica agreed on confirmed status at κ = 0.79 (95% CI 0.78–0.80) (articles 0.71; push comments 0.48; services 0.39–0.72, where nearly every submission is confirmed) and, among 4,893 units confirmed by both, on the domains at κ = 0.63–0.85; medication experience and system navigation met the rule for instrument dependence. Against physician labels for information gap on the earlier 50-unit Taiwanese reference set, the full-text instrument reached κ = 0.57 (95% CI 0.34–0.76), the single-unit reference read 0.72 and the two physicians 0.79.

**Table 4.** The eight friction domains: level, how securely each was measured, and candidate responses.

| **Friction domain** | **Forum articles, % (n/N)** | **Submissions, % (n/N)** | **Articles both vendors confirmed: model; second vendor, %** | **Second-vendor κ** | **Physicians A; B, % of randomly sampled articles** | **Physician–physician κ** | **How securely measured** | **Candidate response to evaluate** |
|---|---|---|---|---|---|---|---|---|
| Information gap | 59.5 (1,426/2,395) | 99.6 (3,092/3,105) | 68.7; 61.7 | 0.85 | 93; 86 | 0.71 | Common under every reader; level reader-dependent (physicians 86%–93% of articles) | Patient-readable explanation of results, symptoms and next steps |
| Emotional burden | 39.0 (935/2,395) | 20.4 (633/3,105) | 47.4; 42.2 | 0.80 | 50; 43 | 0.60 | Forum excess reproduced by second vendor and both physicians | Recognizing and addressing distress alongside disease management |
| Lifestyle burden | 22.2 (532/2,395) | 11.9 (368/3,105) | 28.0; 18.3 | 0.64 | 61; 18 | 0.30 | Forum excess reproduced; level varies between physicians | Practical support for restrictions of diet, work, activity and sleep |
| Medication experience<sup>a</sup> | 24.1 (577/2,395) | 25.0 (777/3,105) | 30.4; 24.0 | 0.63 | 39; 14 | 0.53 | Instrument-dependent (second vendor); descriptive | Medication counselling on side effects and doubts |
| System navigation<sup>a</sup> | 16.3 (391/2,395) | 15.0 (466/3,105) | 20.8; 18.9 | 0.84 | 29; 14 | 0.41 | Instrument-dependent; 9.1% of navigation posts engaged with insurance | Guidance on which service, specialty or hospital to approach |
| Access barrier | 6.4 (154/2,395) | 2.5 (79/3,105) | 8.4; 5.8 | 0.71 | 18; 0 | 0.32 | Rare; physicians disagree | Information on appointments, referral and waiting |
| Caregiver burden | 6.6 (157/2,395) | 1.3 (40/3,105) | 8.2; 8.0 | 0.82 | 25; 11 | 0.60 | Concentrated in family-authored posts; rare overall | Support addressed to the family member helping |
| Cost burden | 2.9 (69/2,395) | 0.6 (18/3,105) | 3.7; 6.4 | 0.66 | 32; 4 | 0.21 | Rare under the model; physicians disagree (32% vs 4%) | Financial counselling; better measurement first |

Model rates are on all model-confirmed units (Table 2). Second vendor: rates on the 1,748 forum articles both vendors confirmed, and κ among the 4,893 units both confirmed. Physicians: each physician’s own labels in the random sample, as a percentage of the sampled forum articles that physician judged patient or caregiver voice; κ between physicians on the 108 units both judged patient or caregiver voice (Table 5). Candidate responses are hypotheses for evaluation, not tested interventions; how often a friction is voiced is not its severity or its priority for patients. Specified after the primary results had been inspected. <sup>a</sup>Instrument-dependent domain: medication experience and system navigation met the rule through its contrast criterion (their services-minus-articles difference became significant under the second vendor; Table S4), not through κ.

The two physicians agreed with each other on patient-voice status at κ = 0.60 (95% CI 0.45–0.74; AC1 0.76), and the instrument agreed with each of them less closely (κ 0.32 [95% CI 0.14–0.49] and 0.37 [0.21–0.53]; Table 5A; every field in Table S6 in Multimedia Appendix 1). On the domain rows, physician–physician κ ranged from 0.21 to 0.71 and instrument–physician κ from 0.15 to 0.67. Of the units the instrument confirmed, the physicians confirmed 97%–99% of submissions, about three quarters of articles (76% and 76%) and about half of push comments (41% and 59%; Table 5B). Conversely, they judged 9 of 15 and 7 of 15 sampled articles the instrument had rejected to be patient or caregiver voice, so the instrument captured an estimated 52% and 58% of physician-identified patient voice among the articles read, against 97%–98% on the services; in this small sample of missed articles, the physicians labelled burden at least as often as in captured ones (any burden 89% and 86% vs 89% and 57%). The physicians identified information-seeking in 93% and 86% of the articles and 91% and 83% of the submissions they confirmed, against 59% and 100% by the instrument, so the need to understand was common under every reader, but its level, and the instrument’s services-minus-articles difference in it (+40.5 points, against −2.2 and −2.5 under the physicians), were reader-dependent. In the random sample, both physicians found more emotional and lifestyle burden in articles than in submissions, as the instrument did on the full sample, and one also found more caregiver and cost burden (Table 5C); both found a larger any-burden difference (−29.2 and −28.4 points, against −9.2 by the instrument on the same sampled units, each source using its own patient-voice judgments). One physician labelled cost burden in 32% of articles (the other 4%; the instrument 3%), lifestyle burden in 61% (18%; 24%) and access barrier in 18% (0%; 0%), and physician–physician κ on these three domains was 0.21–0.32.

**Table 5.**
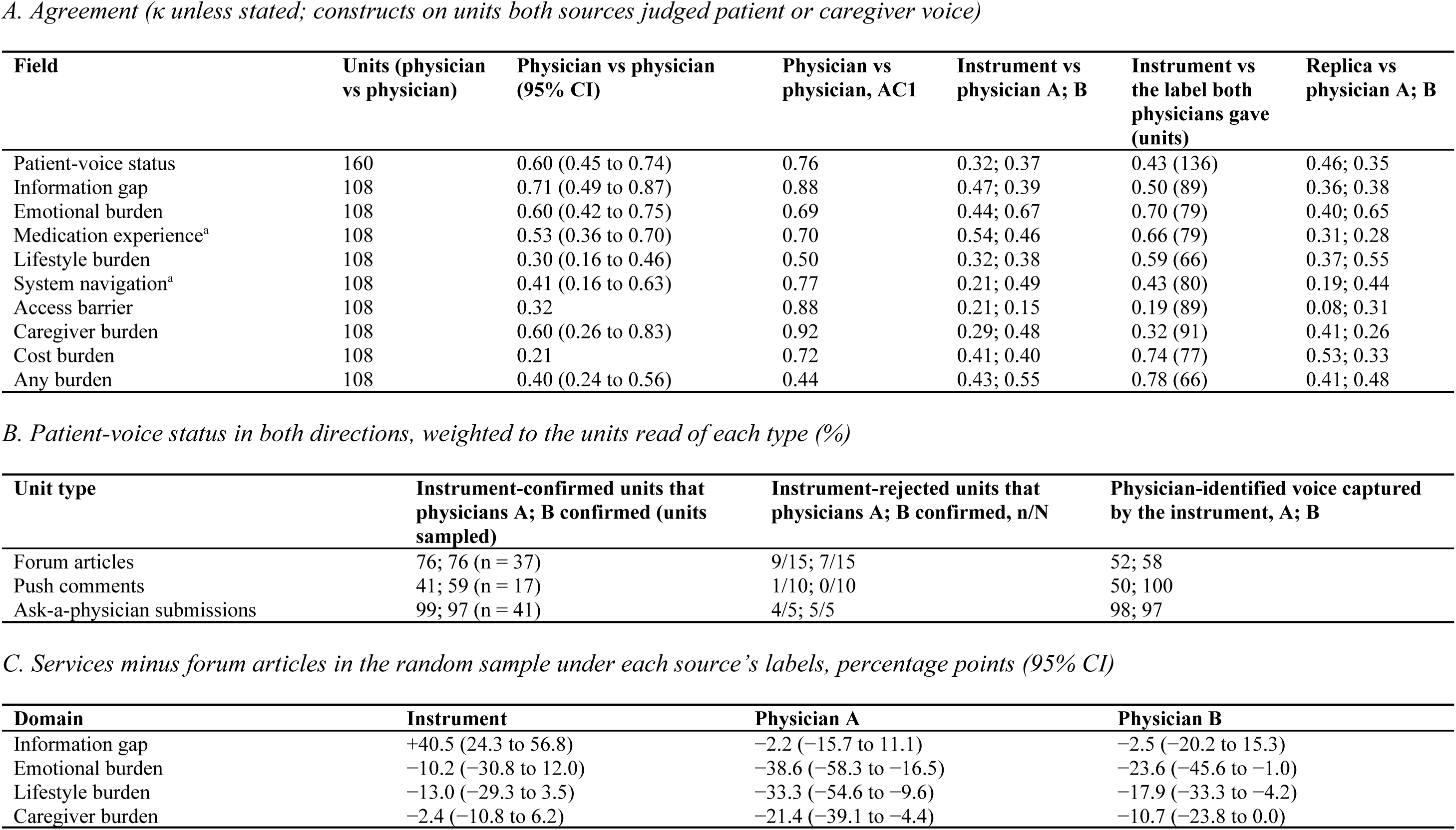
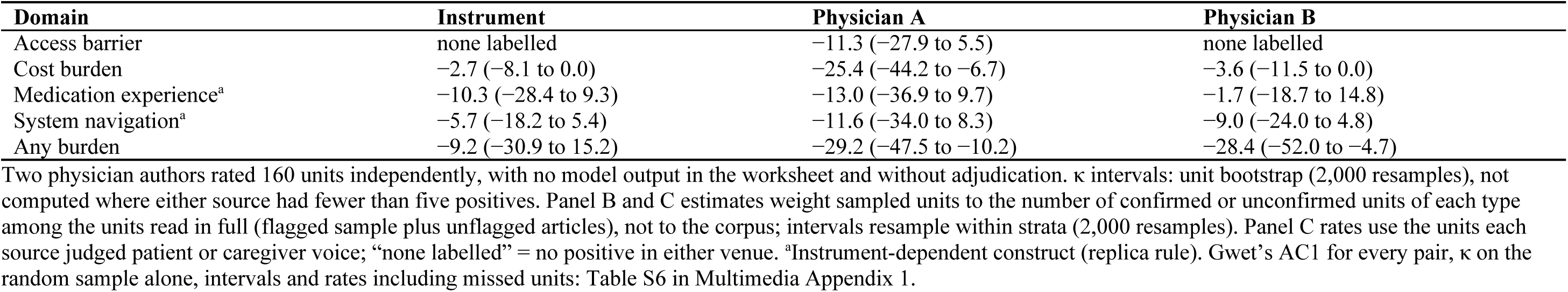
Two physicians’ labels compared with the full-text instrument.

### How Selection Changes the Profile

The friction profile depended on which units were counted. The relevance read confirmed 50.5% (95% CI 49.1–52.0) of flagged articles, 19.7% (95% CI 17.5–22.1) of flagged push comments and 96.1%, 98.2% and 95.4% of KingNet, Taiwan e-Hospital and tcm.tw submissions (Table 1). On flagged rather than confirmed articles, 32.4% voiced a burden and 33.3% a question, against 63.4% and 59.5% of confirmed articles, so on flagged units the forum appeared the less burdened source (any burden 32.4% vs 49.3%–58.7% of flagged submissions on the three services), the reverse of the confirmed picture. The flag also missed patient voice: 244 of 1,000 unflagged articles (24.4%) were confirmed, so the flag reached an estimated 42.9% (95% CI 40.2–45.7) of patient-voice articles, and the archive holds about 225 thousand patient-voice articles (210–239 thousand) rather than the 96 thousand inside the flag. Missed articles were shorter (median 172 vs 482 characters), more often questions (77.9% vs 59.5%) and less often emotional (29.5% vs 39.0%), with similar overall burden (60.2% vs 63.4%); adding them mattered for the information gap more than for burden (33.3% on all flagged, 59.5% on confirmed flagged and 70.0% on all confirmed articles weighted to the archive; any burden 32.4%, 63.4% and 61.6%). These counts refer to the model’s relevance read, which the physicians found conservative on articles; the physicians also rejected about a quarter of model-confirmed articles, so the archive-level direction of bias is uncertain, and both estimates are conditional on the 41 boards in the sample. Applied to the flagged frame, PTT’s 501 thousand flagged units correspond to about 158 thousand model-confirmed units (95% CI 150–165 thousand), so PTT’s share of model-confirmed patient voice in the frame is about 27% rather than 53%. A pooled rate estimates a specified mixture of sources correctly; the problem arises when the mixture is set by the sampling frame: at the projected article share among model-confirmed articles and submissions in the flagged frame (18.5%; the confirmed sample was 43.5% articles because of the oversampling), the pooled value lay outside both source CIs for seven of nine measures, and the pooled information-gap rate moves from 99.6% to 59.5% as the article share moves from 0 to 1 (Figure 3a); the three article denominators are compared in Figure 3b. Of the 2,343 flagged articles not confirmed, the relevance read coded 2,007 as other voices or topics, 258 as media and 78 as clinician voice, and neither removing the physician’s reply from KingNet units nor excluding tcm.tw or KingNet changed any service domain rate by more than 2.1 points.

**Figure 3.**
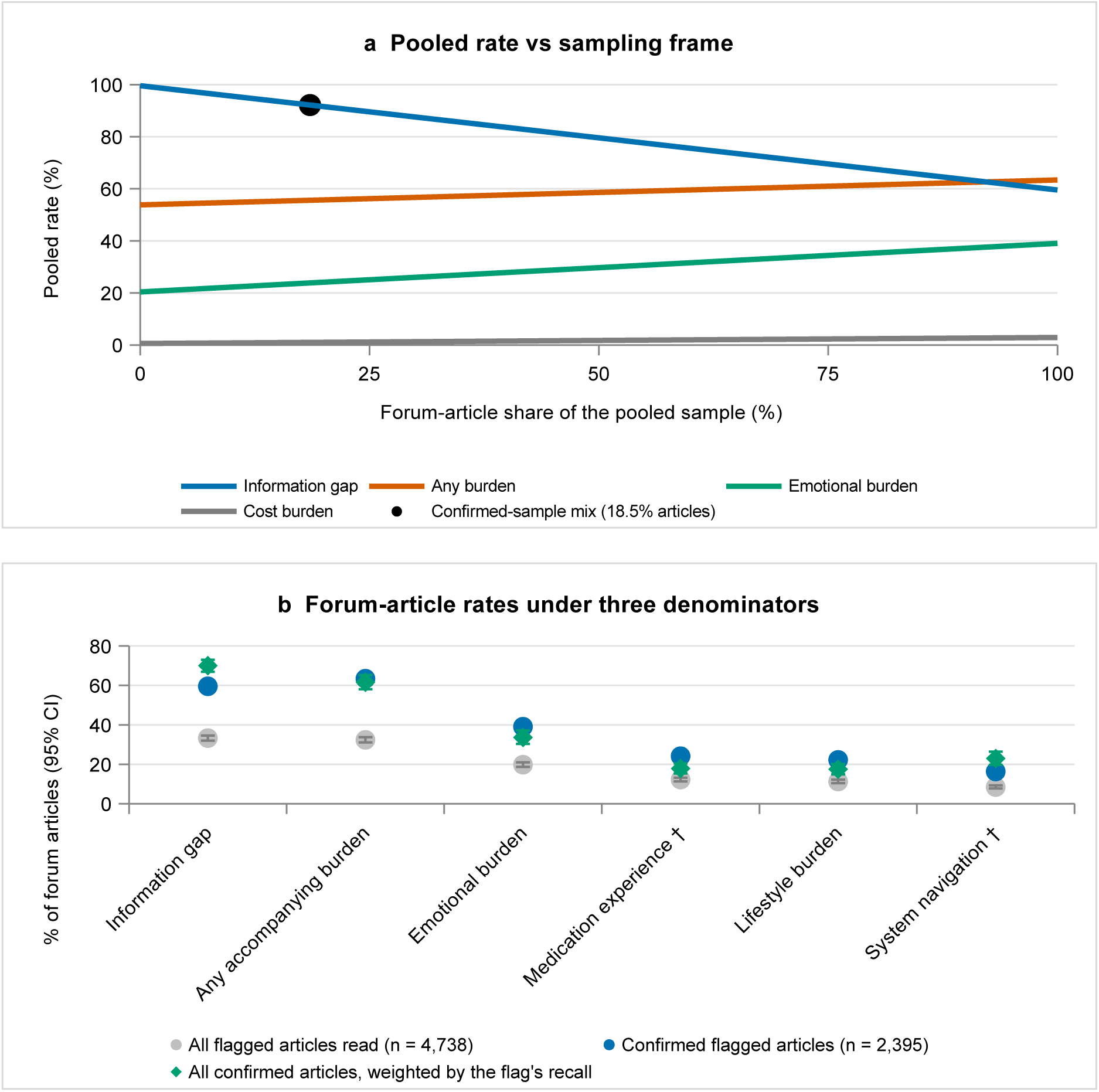
Pooling and the denominator. (a) Pooled rate as a function of article share for four constructs (information gap, any burden, emotional burden and cost burden), with each source’s 95% CI as a shaded bar at 0% and 100% and the information-gap value marked at the projected article share among model-confirmed articles and submissions in the flagged frame (18.5%); the curves illustrate dependence on composition, not a target population estimate. (b) Six constructs on forum articles under three denominators: all flagged articles read (unconfirmed), confirmed flagged articles, and all confirmed articles weighted to the archive through the flag’s recall (stratified bootstrap 95% CI).

### Keyword Classifier Benchmark

The production pipeline’s information-gap output had been computed for only 680 of 2,272 model-confirmed units on the reference stratum (29.9%): 37 of 382 forum articles (9.7%, mostly recent posts), 630 of 699 KingNet submissions, 13 of 234 push comments and none of the Taiwan e-Hospital or tcm.tw submissions (Table S7 in Multimedia Appendix 1; Figure 4b; the flag’s precision and recall are in Figure 4a). As stored, with missing outputs counted as absent, the pipeline showed information-seeking in 5.8% of forum articles against 58.6% by the full-text model, a 10.2-fold difference that reflects missing output rather than lexical error. Where the output had been computed, the keyword and model rates were similar on forum articles (59.5% vs 51.4%; n = 37) and lower on KingNet (67.5% vs 100.0%). The NHI lexicon, applied to every unit, over-counted NHI navigation 2.9-fold relative to the strict model read (4.2% vs 1.5%; precision 30.5%; Figure 4c). A friction map built from the stored pipeline outputs would have presented the need to understand as a minority concern, mainly because the output was missing, and would have overestimated insurance navigation.

**Figure 4.**
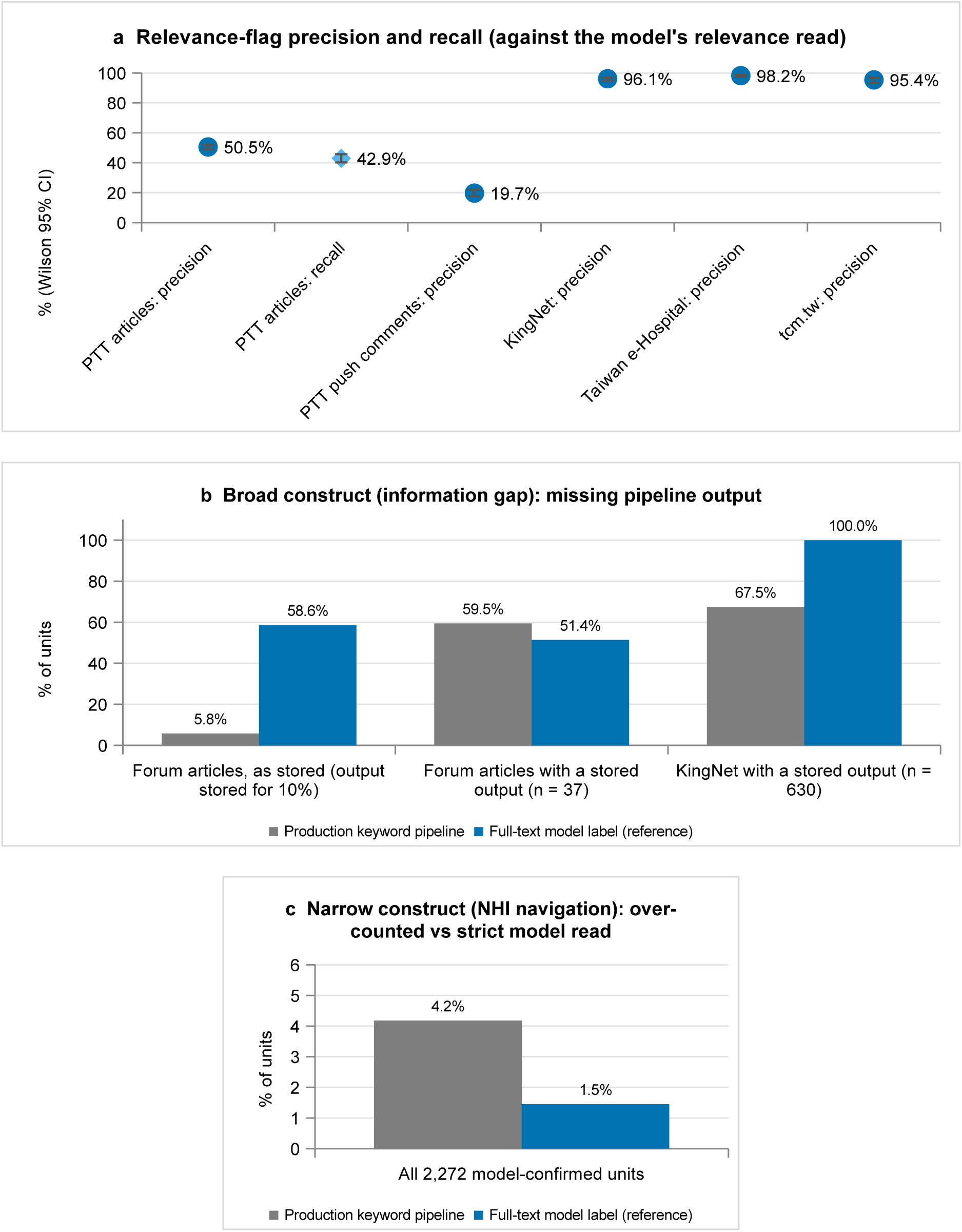
Measurement quality. (a) Precision of the production patient-voice flag by unit type (share of flagged units confirmed by the model’s relevance read, Wilson 95% CI) and its recall on forum articles (open diamond), both model-referenced. (b) Broad construct: the pipeline’s information-gap output as stored (missing counted as absent) and on units where it had been computed, against the full-text model label; Taiwan e-Hospital and tcm.tw had no stored output. (c) Narrow construct: NHI navigation over-counted relative to a strict model read. Rates, coverage and fold differences are in Table S7 in Multimedia Appendix 1.

## Discussion

### Principal Findings

Reading the full text of a Taiwanese sample under an explicit eight-domain framework showed what patients and families bring to public channels. The need to understand (is this normal, what does this result mean, what should I do?) led under every reader, and emotional burden followed under the model and the second vendor; medication experience, lifestyle restriction and navigation were common, and access barriers, caregiver burden and explicit financial strain were voiced less often. Family members wrote one post in seven, and their forum posts more often voiced distress, navigation and access difficulty, with their own strain usually accompanied by distress, whereas patients’ posts more often voiced daily-life restriction. Distress came with questions as often as without them, while some frictions clustered: distress with lifestyle restriction, navigation with access difficulty, and a relative’s strain with distress. The forum carried more emotional and lifestyle burden than physician submissions on every reading, and more family burden under the model, and a physician submission was almost always a question, so the two sources show different parts of the same profile.

Domains differed in how securely they were measured. Emotional burden and the forum’s excess of emotional and lifestyle burden were reproduced by a second vendor and by both physicians; the need to understand was common under every reader but its level was reader-dependent (59.5% of forum articles by the model, 86%–93% by the physicians); medication experience and navigation were instrument-dependent; and cost and access burden were rare under the model and disputed between the physicians. Lower agreement on a domain limits its exact rate, not its relevance: a friction that two physicians label differently is still a friction patients express, and Table 4 states for each domain what can and cannot be claimed.

The profile was also sensitive to how units were selected. With 50.5% precision on articles and 19.7% on comments, the pipeline’s flagged denominator diluted the forum’s friction rates and reversed the overall source comparison, and with 42.9% recall the flag missed shorter, question-dominated posts. The production pipeline’s stored outputs omitted the dominant friction for most units and its insurance lexicon over-counted a rare institutional one about threefold. Frictions that patients do express can therefore disappear, or be distorted, in a keyword-based summary.

### Comparison With Prior Work

Platform features shape patient-generated content [8,9,32,34,35]; this study adds a multi-domain friction profile for Traditional Chinese patient voice, with authorship and measurement security stated for each domain. Our finding that emotional content concentrates on a peer platform agrees with the Dutch comparison of platforms [32] (39.0% vs 20.4% in confirmed units), although our forum is a general-purpose board in a single-payer setting. Against the patient-listening guideline [6], the study implements collection, selection and coding and reports counts at each step (Table 1); emotional and lifestyle burden map onto its quality-of-life facets and medication experience onto treatment experience, while the other domains add the health-system and informational concepts of unmet need and burden of living with a condition [5,7]. Two departures are candidates for its next revision: on a general-purpose forum a keyword relevance flag does not yield a patient cohort, and family members writing for patients should be distinguished from patients, because they voice different frictions.

LLM annotation of health social-media text has been validated against human labels [25–28] and keyword over- and under-counting are well documented [20,21]. Here the instrument agreed with the physicians less than they agreed with each other on relevance and on information-seeking, and its relevance read was conservative on forum articles, which a check of confirmed units alone would not have revealed.

### Implications

The frictions point to candidate responses that can be evaluated (Table 4). The most frequent need, understanding results, symptoms and next steps, points to patient-readable explanation delivered where patients already look. Two in five forum questions came with distress, so information alone may not be enough. Navigation rarely concerned insurance, which suggests that guidance on where to seek care matters in a system without gatekeeping. Family-authored posts more often voiced navigation and access difficulty and their own strain, which argues for information and support addressed to the person helping, while respecting patients’ own priorities. These are hypotheses for communication and service design: frequency is not severity or patient priority, an expressed concern is not proof of an unmet need, and none of these responses has been tested here.

For patient-focused drug development and surveillance, a friction framework with explicit definitions, author coding and per-domain measurement checks is transferable; the source and unit type are reporting units; and a relevance classifier’s precision and recall, and whether a keyword pipeline stored a label at all, must be measured before its counts are trusted. For LLM-based listening, the transferable elements are a full-text relevance read, verbatim rubrics, an agreement chain to an independent read and physician labels, a physician check of both accepted and rejected units, a second-vendor replica with a pre-specified rule, and published prompts.

### Limitations

First, the framework was defined by the investigators from existing concepts, not derived from these posts; frictions outside the eight domains were not measured. Patients and caregivers did not help define the domains [5,6]. Second, the physician comparison was modest: two physician authors, aware of the study aims and one of whom designed the instrument, rated 160 units without adjudication; they agreed with each other only moderately (κ 0.21–0.71 across domains; 0.60 on patient-voice status, where the instrument’s agreement with each physician, 0.32 and 0.37, was lower), and one labelled several domains far more often than the other. Third, relevance was confirmed by the model, and the physicians judged that it captured about half of the patient voice in the forum articles read (52% and 58%), estimated from only 15 rejected articles; a revised, recall-oriented relevance rubric, frozen (SHA-256) before use, was developed afterwards for a companion study (Liu ZY, Hung YC, Kuo CF; preprint submitted to medRxiv), and the rates here are not revised retrospectively.

Fourth, the information-gap domain combines an explicit request for information, expressed uncertainty and the question format of a consultation; a future codebook should separate these, and medication experience and navigation, which were instrument-dependent and can be neutral, should be split into neutral and burdensome subtypes. Fifth, rates describe what self-selected posters on four platforms expressed, not population prevalence, and how often a friction is voiced is not its severity, its importance to patients or evidence that care missed it; showing that such concerns are omitted from encounters or records would need a comparison with those records. Sixth, recall was measured on forum articles of the sampled boards only. Seventh, the archive spans 1995–2026; restriction to 2020 or later preserved the source contrast (−9.4 points, 95% CI −15.1 to −3.7), and author-clustered intervals were close to unit-level ones (width ratios 0.96–1.08; the 2020 restriction is in Table S4 in Multimedia Appendix 1). Eighth, the comparisons of patient-with family-authored posts and the co-occurrence summaries were specified after the primary results had been inspected and are descriptive. Ninth, sources differ in their authors, topics and eras (median year 2015 vs 2012), the rates come from one language and one pipeline, and the models may have seen public PTT posts during training. Finally, the companion study draws on the same archive (338 of the 36,564 distinct posts in its seven condition frames are among the units read here) but applies the framework to kidney-related posts and reports none of the results presented here.

## Conclusions

Systematic listening to Taiwanese online health discussions, read in full under an explicit, Taiwan-contextualized framework, organized patients’ and families’ accounts into eight friction domains. The need to understand and emotional burden led; navigation rarely concerned insurance; family-authored posts more often voiced navigation, access difficulty and distress; and the forum carried more emotional and daily-life burden than physician submissions under every reader. These are concerns that may not surface in routine encounters and are candidates for patient-centred communication and services, which require evaluation. Their exact levels depended for some domains on the reader, and keyword pipelines missed or distorted them, so listening of this kind needs full-text reading, explicit definitions, author coding, unit-stratified denominators with measured precision and recall, and clinician checks of both accepted and rejected content.

## Acknowledgments

We thank the patient communities whose public posts make this research possible.

## Generative AI Disclosure

Generative artificial intelligence was used in two distinct roles. As study instruments, gpt-6.1-sol (OpenAI; Codex command-line interface) produced the full-text relevance and construct annotations; Claude Sonnet 5.5 (Anthropic) produced the second-vendor replica; GPT-5.5 (OpenAI; OpenRouter route openai/gpt-5.5) produced the single-unit reference read and the strict NHI-navigation read on the reference stratum; and gpt-5.6-sol and Claude Opus 5.5 took part in the instrument pilot. This use, including prompts, batch sizes and settings, is specified in the Methods and Multimedia Appendix 2. In manuscript preparation, Anthropic Claude (Claude Code and Cowork; Claude Opus models including Claude Opus 5.5) assisted with analysis code, drafting and editing under the authors’ direction; the authors reviewed and verified the code and every number and statement, and take full responsibility for the content.

## Data Availability

The data supporting this study are available from the corresponding author on reasonable request. Post text is not shared, because the platforms’ terms prohibit redistribution of user content.

## Authors’ Contributions

Contributor roles (CRediT): CFK, conceptualization, methodology, formal analysis, investigation (physician comparison), data curation, visualization, writing – original draft, writing – review and editing, supervision; ZYL, data curation, resources, writing – review and editing; CT, investigation (physician comparison), writing – review and editing. CFK had full access to all data and takes responsibility for the integrity of the data and the accuracy of the analysis. All authors approved the final version and agreed to its submission and posting.

## Abbreviations

AC1: Gwet first-order agreement coefficient
CI: confidence interval
LLM: large language model
MOHW: Ministry of Health and Welfare
NHI: National Health Insurance
pp: percentage points
SHA-256: Secure Hash Algorithm 256-bit
STROBE: Strengthening the Reporting of Observational Studies in Epidemiology
TRIPOD-LLM: Transparent Reporting of a Multivariable Model for Individual Prognosis or Diagnosis–Large Language Models

## Supplementary Material

The supplementary material follows as three Multimedia Appendices.

### Multimedia Appendix 1. Tables S1–S7

Tables S1–S7 support the main text: platform terms audit (S1), per-site rates on the three services (S2), sample characteristics (S3), robustness of the source contrast (S4), agreement with the second vendor, the single-unit reference read and earlier physician reference sets (S5), the two-physician comparison (S6) and the keyword pipeline benchmark (S7).

**Table S1.** Platform terms audit (live, 2026-08-16)

| **Platform** | **Verdict** | **Operative clause (verbatim snapshot archived)** |
|---|---|---|
| Taiwan e-Hospital | Included | MOHW Government Website Open Data Declaration: free, non-exclusive, sublicensable licence incl. reproduction/adaptation; attribution required |
| PTT | Included (conditions) | No term or robots.txt (HTTP 404) restricting automated access; §1-2 user licence is to PTT (non-profit) and does not extend to third-party republication → no verbatim reproduction; over18 age-gate cookie disclosed |
| KingNet | Included (conditions) | Terms bar commercial use and excerpting; robots.txt permits general crawl (Crawl-delay 5) outside /admin, /ajax/*; non-commercial aggregate use only |
| tcm.tw | Included (conditions) | Footer bars re-publication; robots.txt restricts only bingbot on Q&A views; aggregate, non-republishing analysis |
| 5914 | Excluded | robots.txt: User-agent: * / Disallow: / — all non-search agents disallowed site-wide |
| Dcard | Excluded | Terms (2025-04-24) prohibit automated collection of user content and circumvention of access-limiting measures |
| BabyHome | Excluded | Terms bar unauthorized use “in any form, for any purpose”; robots.txt disallows generic crawlers; Content-Signal ai-train=no |
| Patient-association sites | Excluded | Terms unverifiable at audit date |

**Table S2.** Per-site construct rates on the three ask-a-physician services and heterogeneity (full-text annotation)

| Construct | KingNet (n = 1,318), % (95% CI) | Taiwan e-Hospital (n = 1,293) | tcm.tw (n = 494) | Range, pp | χ² P (Holm across the 7 burdens) |
|---|---|---|---|---|---|
| Information gap (question) | 99.9 (99.6–100.0) | 99.5 (98.9–99.7) | 99.0 (97.7–99.6) | 0.9 | .02 |
| Emotional burden | 19.4 (17.4–21.6) | 22.8 (20.6–25.2) | 16.6 (13.6–20.1) | 6.2 | .04 |
| Medication experience | 24.1 (21.9–26.5) | 25.1 (22.8–27.6) | 27.1 (23.4–31.2) | 3.0 | .92 |
| Lifestyle burden | 11.8 (10.2–13.7) | 10.6 (9.0–12.4) | 15.2 (12.3–18.6) | 4.6 | .14 |
| System navigation | 13.0 (11.3–14.9) | 12.8 (11.1–14.8) | 26.1 (22.4–30.2) | 13.3 | <.001 |
| Access barrier | 2.6 (1.9–3.6) | 2.2 (1.5–3.1) | 3.4 (2.2–5.4) | 1.3 | .92 |
| Caregiver burden | 1.8 (1.2–2.7) | 0.9 (0.5–1.6) | 0.8 (0.3–2.1) | 1.0 | .30 |
| Cost burden | 0.7 (0.4–1.3) | 0.4 (0.2–0.9) | 0.8 (0.3–2.1) | 0.4 | .92 |
| Any burden other than a question | 51.1 (48.4–53.8) | 53.8 (51.1–56.5) | 60.9 (56.6–65.1) | 9.8 | <.001 |

Design weights (flagged population ÷ units read): KingNet 136.8, Taiwan e-Hospital 137.2, tcm.tw 135.3; the largest weighted-minus-unweighted gap on any construct was 0.02 percentage points.

**Table S3.**
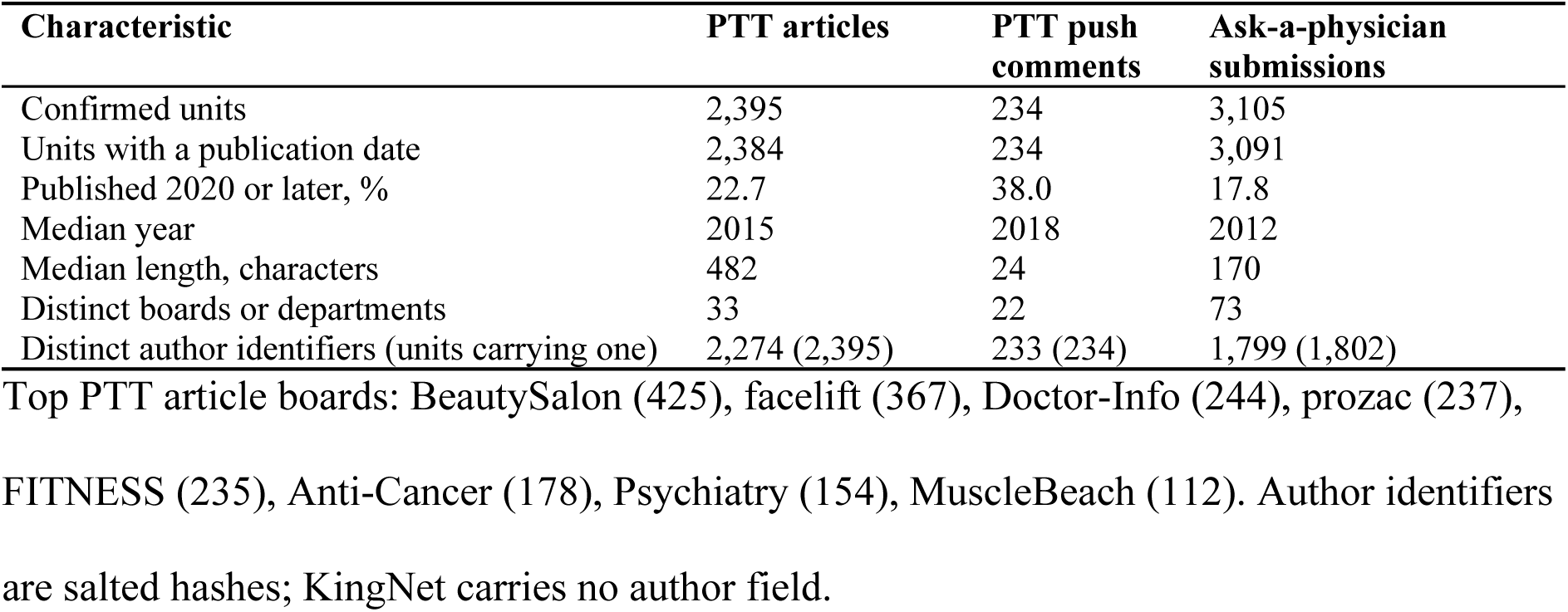
Sample characteristics of confirmed units by unit type.

**Table S4.** Robustness of the source contrast: services minus forum articles, percentage points (95% CI)

| Analysis | Articles / submissions, n | Emotional | Lifestyle | Caregiver | Access | Cost | Medication † | Navigation † | Any burden |
|---|---|---|---|---|---|---|---|---|---|
| Primary: all confirmed units (main-text Table 2) | 2,395 / 3,105 | −18.7 (−21.1 to −16.2)* | −10.4 (−12.4 to −8.4)* | −5.3 (−6.4 to −4.2)* | −3.9 (−5.1 to −2.8)* | −2.3 (−3.1 to −1.6)* | +0.9 (−1.4 to 3.2) | −1.3 (−3.3 to 0.6) | −9.6 (−12.2 to −6.9)* |
| Random stratum of the flagged frame only | 382 / 3,105 | −13.6 (−18.7 to −8.8)* | −8.8 (−13.3 to −4.9)* | −3.9 (−6.7 to −2.1)* | −3.7 (−6.7 to −1.6)* | −2.0 (−4.2 to −0.8)* | +4.9 (0.3 to 8.9) | +0.3 (−3.7 to 3.8) | −4.8 (−10.0 to 0.5) |
| Board-restricted oversample only | 2,013 / 3,105 | −19.6 (−22.2 to −17.0)* | −10.7 (−12.8 to −8.5)* | −5.5 (−6.8 to −4.4)* | −3.9 (−5.2 to −2.7)* | −2.4 (−3.2 to −1.6)* | +0.2 (−2.3 to 2.6) | −1.6 (−3.7 to 0.4) | −10.5 (−13.2 to −7.7)* |
| Within common length tertiles (≤168, ≤386, longer characters; Mantel–Haenszel) | 2,395 / 3,105 | −9.5 (−12.4 to −6.6)* | −4.3 (−6.7 to −2.0)* | −3.4 (−4.6 to −2.2)* | −2.3 (−3.6 to −1.0)* | −2.0 (−2.9 to −1.2)* | +8.6 (5.9 to 11.3)* | −2.3 (−4.7 to 0.1) | −2.4 (−5.4 to 0.6) |
| Appearance and fitness boards excluded | 1,210 / 3,105 | −34.2 (−37.4 to −31.1)* | −17.0 (−19.8 to −14.2)* | −11.4 (−13.5 to −9.6)* | −6.5 (−8.4 to −4.9)* | −3.1 (−4.4 to −2.2)* | −6.3 (−9.4 to −3.3)* | −8.9 (−11.6 to −6.2)* | −26.7 (−29.4 to −23.8)* |
| Two-of-three consensus labels (reference stratum) | 375 / 1,659 | −13.9 (−19.2 to −8.8)* | −7.3 (−11.8 to −3.4)* | −3.5 (−6.1 to −1.8)* | −3.2 (−6.1 to −1.1)* | −2.0 (−4.2 to −0.7)* | +2.3 (−2.6 to 6.7) | −0.5 (−4.6 to 3.1) | −5.0 (−10.5 to 0.6) |
| Second-vendor replica (end to end) | replica-confirmed | −19.8 (−22.5 to −17.1)* | −9.4 (−11.4 to −7.3)* | −7.0 (−8.3 to −5.7)* | −3.6 (−4.9 to −2.5)* | −5.4 (−6.7 to −4.3)* | −7.1 (−9.5 to −4.7)* | −5.7 (−7.9 to −3.6)* | −24.1 (−26.7 to −21.3)* |
| Units published in 2020 or later | — | −24.0 (−29.2 to −18.5)* | −12.0 (−16.4 to −7.6)* | −5.7 (−8.2 to −3.6)* | −4.7 (−7.4 to −2.1)* | −2.2 (−3.9 to −0.8)* | +3.0 (−2.3 to 8.3) | −3.0 (−7.1 to 1.2) | −9.4 (−15.1 to −3.7)* |

\* Holm-significant within the row (8 contrasts). The random-stratum row compares the random-stratum articles with all confirmed submissions. Newcombe intervals, except Mantel–Haenszel (Sato) intervals across length tertiles. Appearance and fitness boards: BeautySalon, FITNESS, MuscleBeach, facelift, fatworld, teeth\_salon. Consensus: a unit was confirmed when at least two of the full-text instrument, the replica and the single-unit reference read confirmed it, and a construct was positive when most of the instruments that confirmed the unit labelled it. Topic-matched psychiatry comparison (PTT prozac and Psychiatry boards, n = 391, vs KingNet psychiatry submissions, n = 52): any burden −4.5 (−16.6 to 3.0), emotional −18.1 (−32.2 to −5.7)\*, medication −0.8 (−13.0 to 13.5), lifestyle −3.7 (−16.7 to 10.7) (Holm across 4). Any burden by length tertile, articles vs submissions: short 60.4% vs 39.9%; medium 66.8% vs 65.2%; long 62.4% vs 73.4%. † Instrument-dependent construct.

**Table S5.**
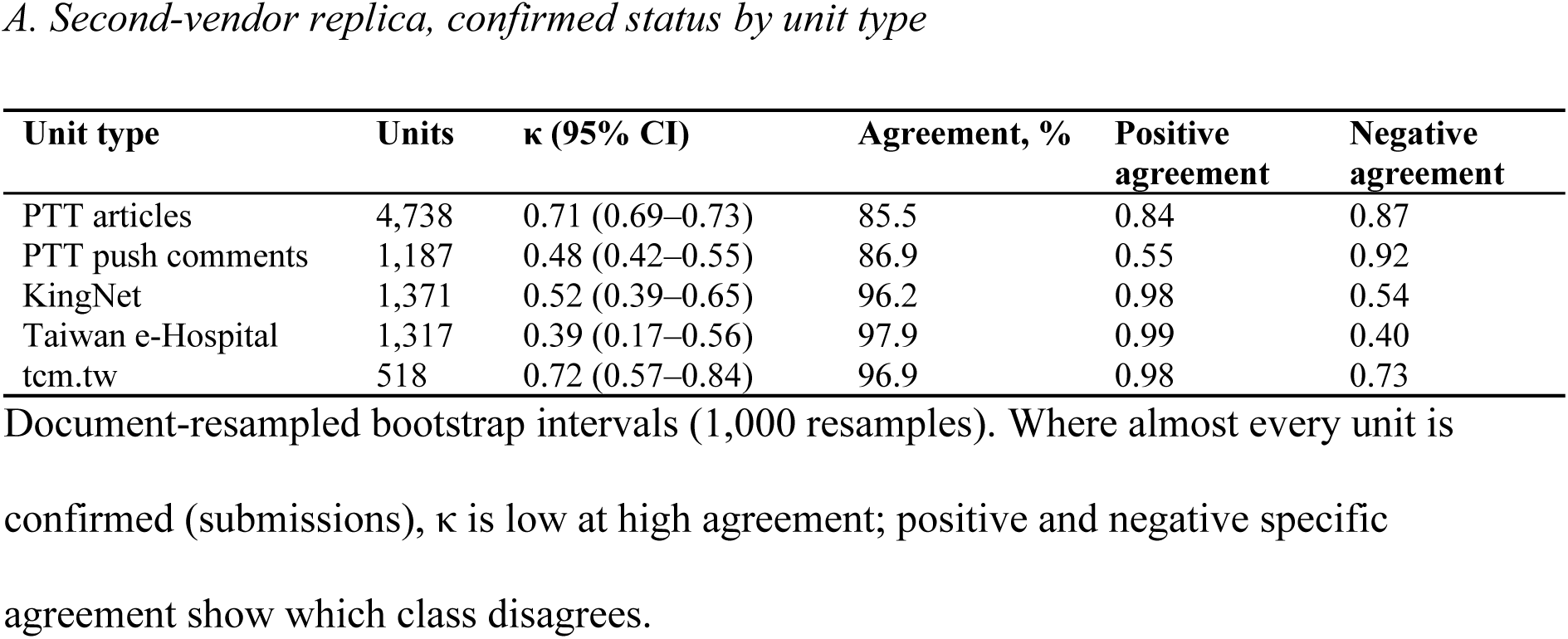
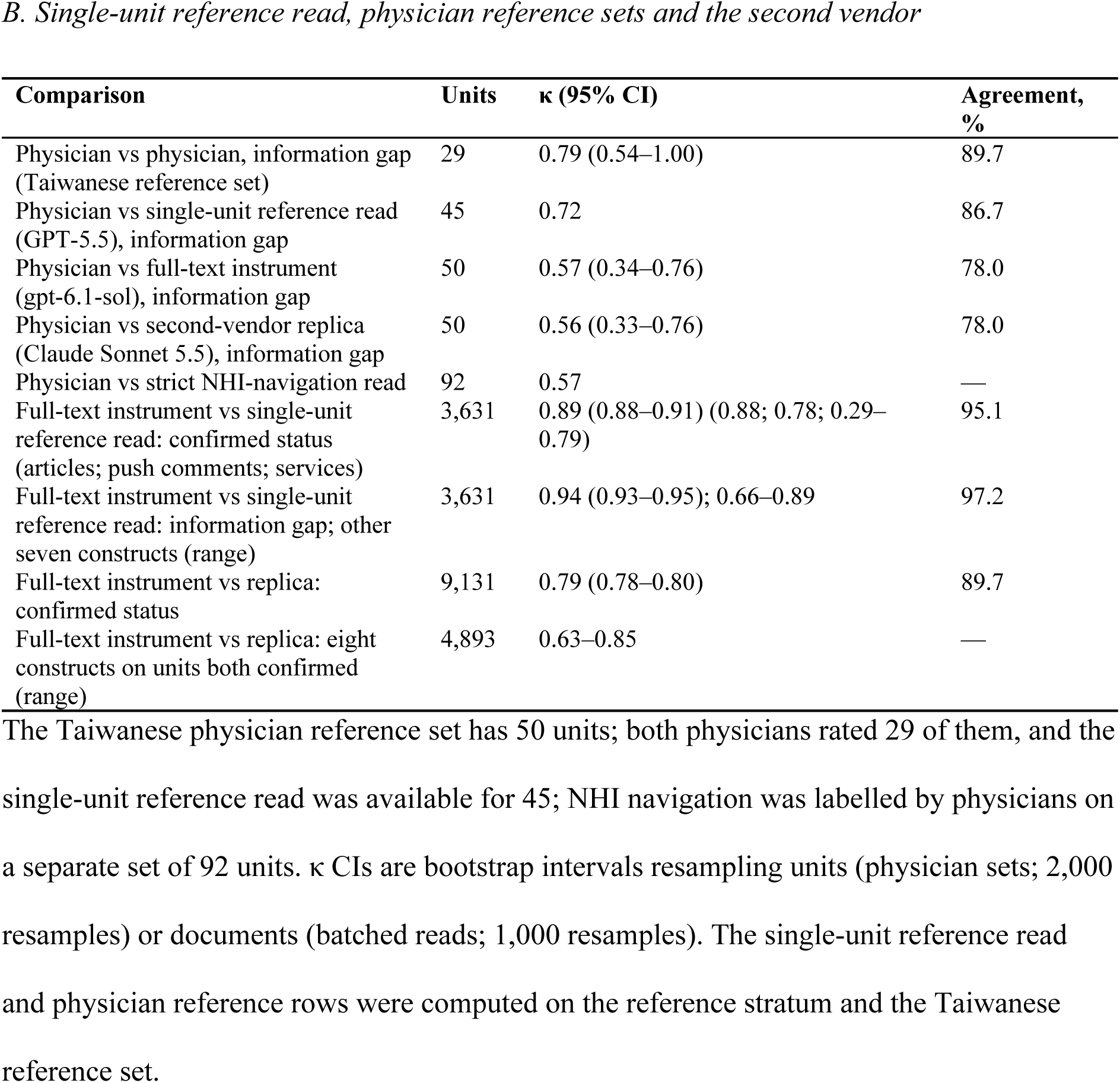
Agreement of the full-text instrument with other readers.

**Table S6.**
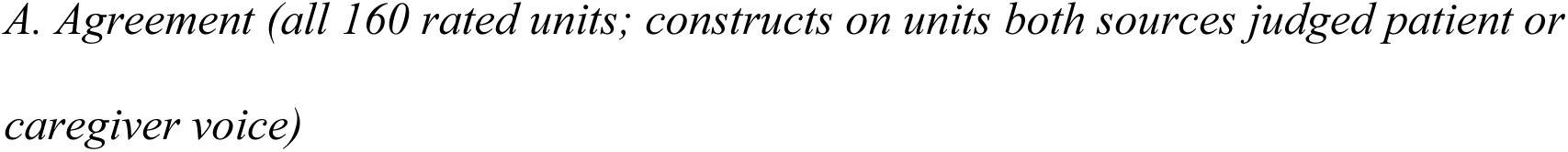
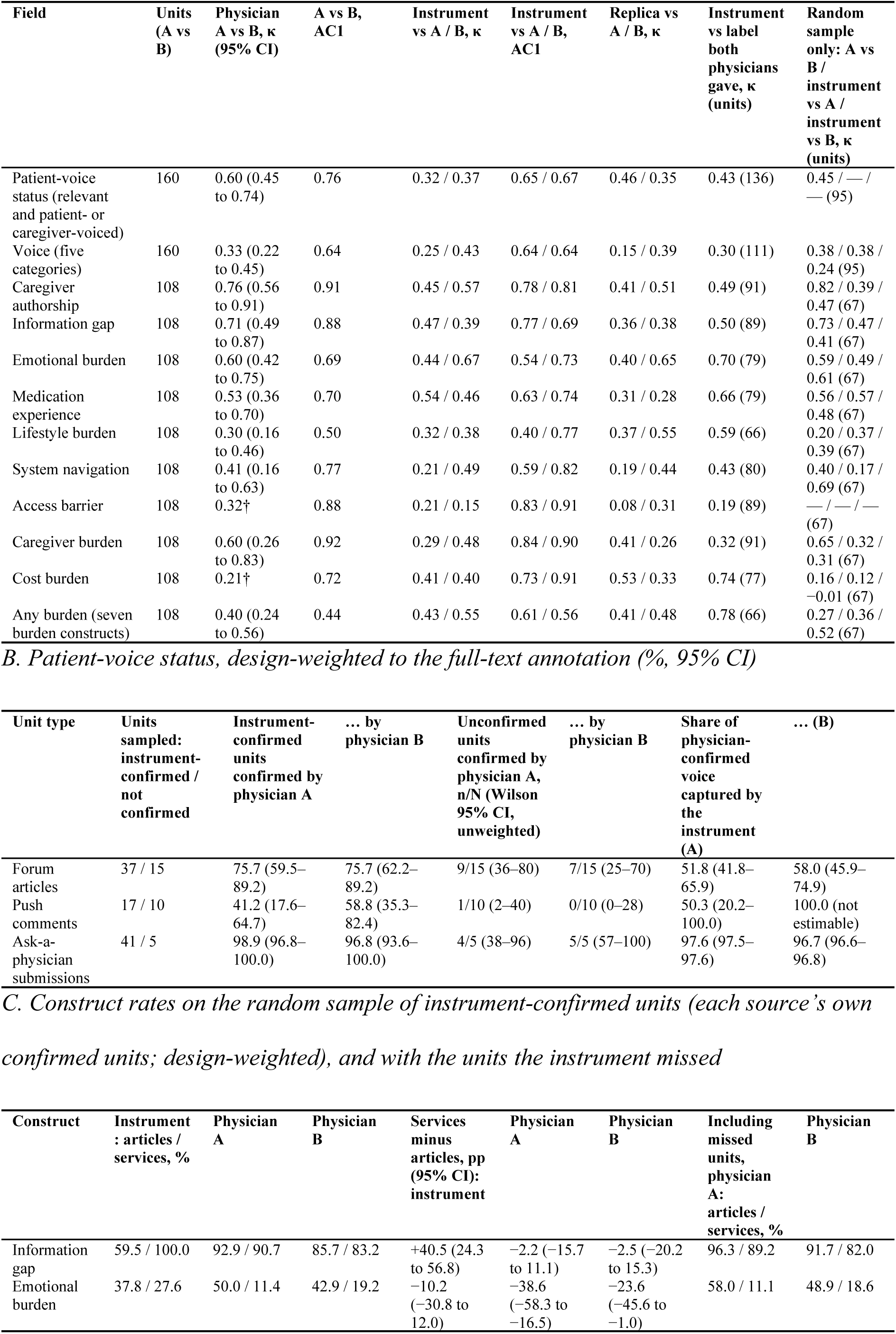
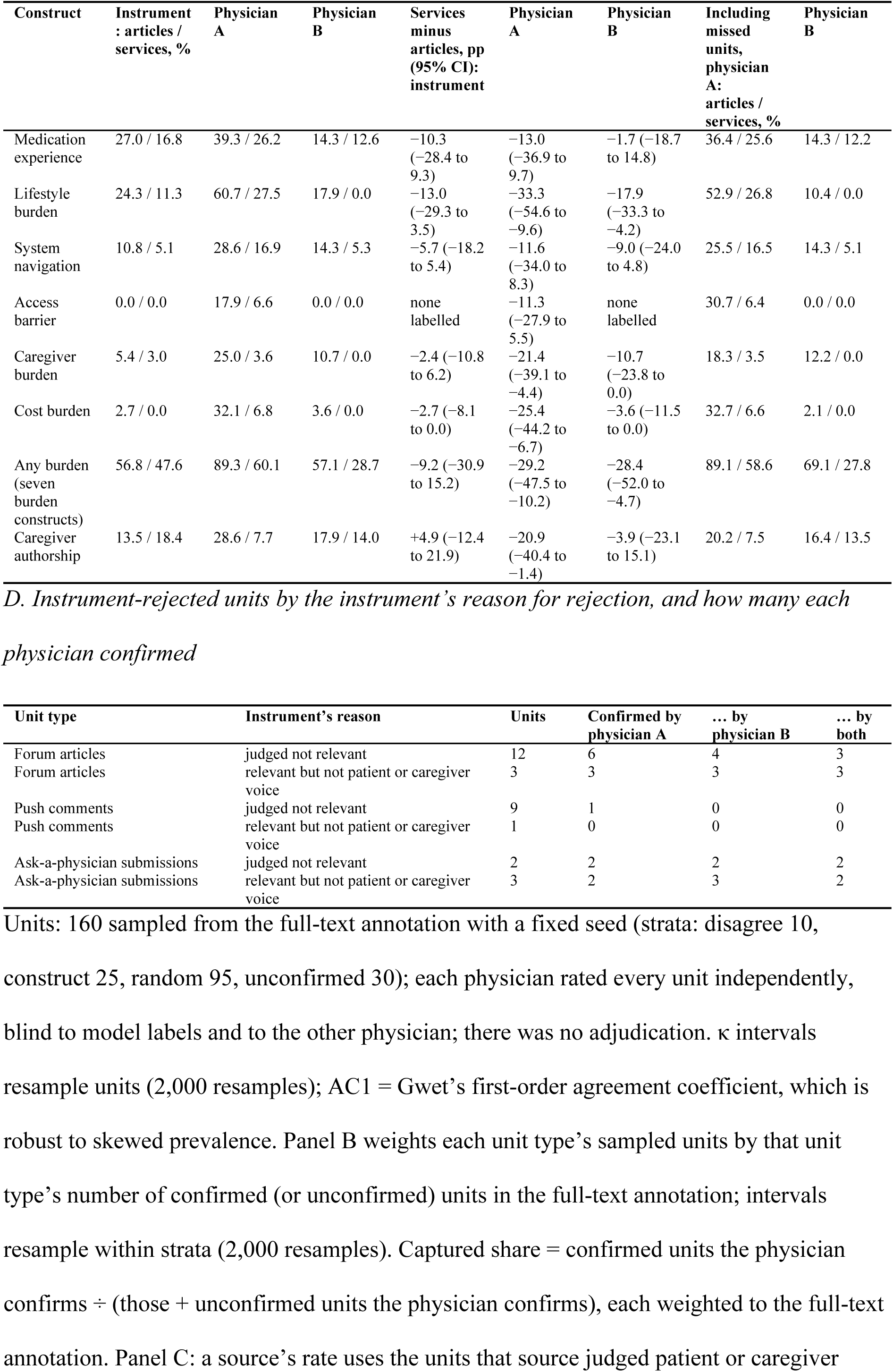
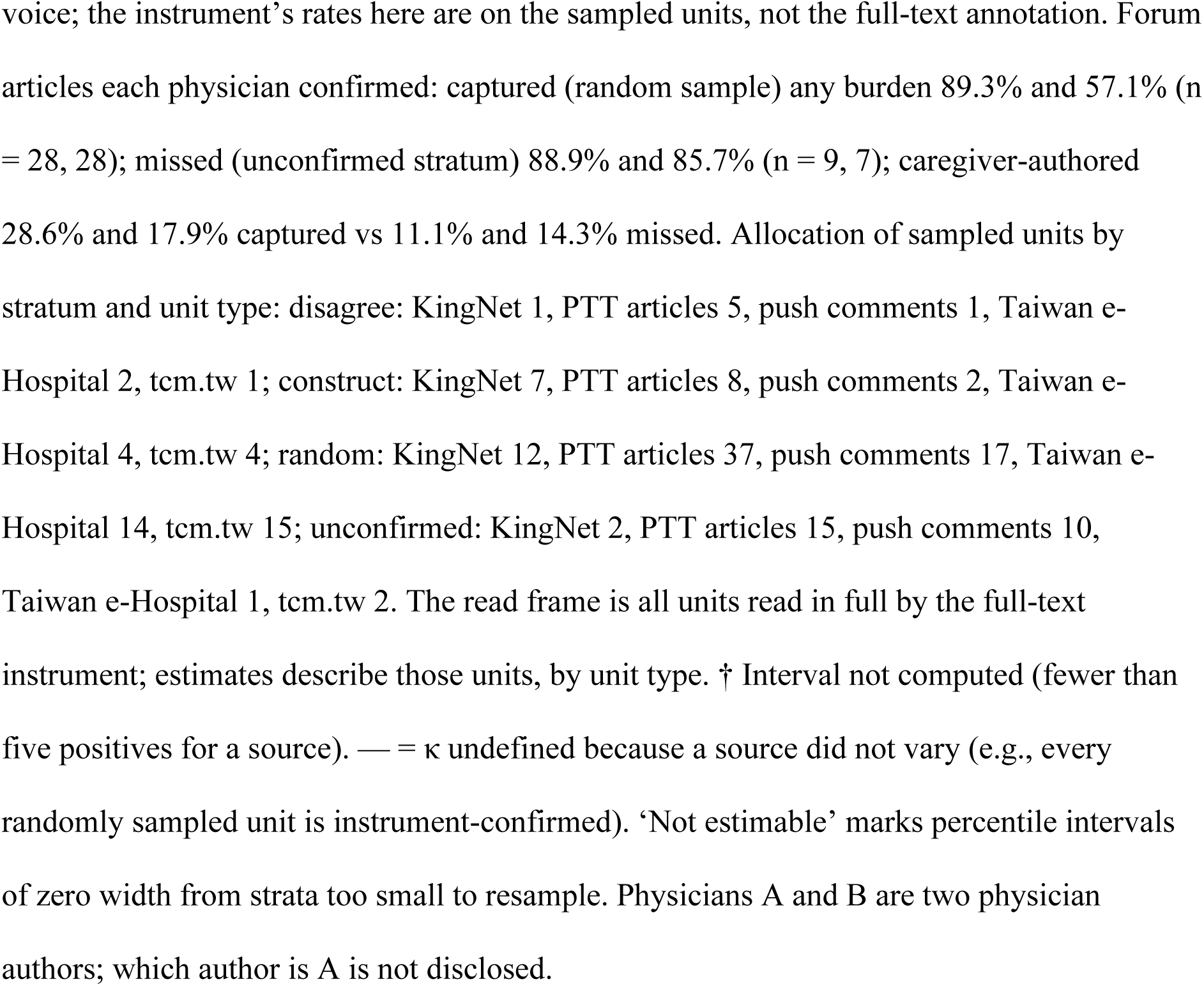
Two-physician comparison with the full-text instrument: agreement, patient-voice status and construct rates.

| *A. Agreement (all 160 rated units; constructs on units both sources judged patient or caregiver voice)* |  |  |  |  |  |  |  |  |
| **Field** | **Units (A vs B)** | **Physician A vs B, κ (95% CI)** | **A vs B, AC1** | **Instrument vs A / B, κ** | **Instrument vs A / B, AC1** | **Replica vs A / B, κ** | **Instrument vs label both physicians gave, κ (units)** | **Random sample only: A vs B / instrument vs A / instrument vs B, κ (units)** |
|---|---|---|---|---|---|---|---|---|
| Patient-voice status (relevant and patient- or caregiver-voiced) | 160 | 0.60 (0.45 to 0.74) | 0.76 | 0.32 / 0.37 | 0.65 / 0.67 | 0.46 / 0.35 | 0.43 (136) | 0.45 / — / — (95) |
| Voice (five categories) | 160 | 0.33 (0.22 to 0.45) | 0.64 | 0.25 / 0.43 | 0.64 / 0.64 | 0.15 / 0.39 | 0.30 (111) | 0.38 / 0.38 / 0.24 (95) |
| Caregiver authorship | 108 | 0.76 (0.56 to 0.91) | 0.91 | 0.45 / 0.57 | 0.78 / 0.81 | 0.41 / 0.51 | 0.49 (91) | 0.82 / 0.39 / 0.47 (67) |
| Information gap | 108 | 0.71 (0.49 to 0.87) | 0.88 | 0.47 / 0.39 | 0.77 / 0.69 | 0.36 / 0.38 | 0.50 (89) | 0.73 / 0.47 / 0.41 (67) |
| Emotional burden | 108 | 0.60 (0.42 to 0.75) | 0.69 | 0.44 / 0.67 | 0.54 / 0.73 | 0.40 / 0.65 | 0.70 (79) | 0.59 / 0.49 / 0.61 (67) |
| Medication experience | 108 | 0.53 (0.36 to 0.70) | 0.70 | 0.54 / 0.46 | 0.63 / 0.74 | 0.31 / 0.28 | 0.66 (79) | 0.56 / 0.57 / 0.48 (67) |
| Lifestyle burden | 108 | 0.30 (0.16 to 0.46) | 0.50 | 0.32 / 0.38 | 0.40 / 0.77 | 0.37 / 0.55 | 0.59 (66) | 0.20 / 0.37 / 0.39 (67) |
| System navigation | 108 | 0.41 (0.16 to 0.63) | 0.77 | 0.21 / 0.49 | 0.59 / 0.82 | 0.19 / 0.44 | 0.43 (80) | 0.40 / 0.17 / 0.69 (67) |
| Access barrier | 108 | 0.32† | 0.88 | 0.21 / 0.15 | 0.83 / 0.91 | 0.08 / 0.31 | 0.19 (89) | — / — / — (67) |
| Caregiver burden | 108 | 0.60 (0.26 to 0.83) | 0.92 | 0.29 / 0.48 | 0.84 / 0.90 | 0.41 / 0.26 | 0.32 (91) | 0.65 / 0.32 / 0.31 (67) |
| Cost burden | 108 | 0.21† | 0.72 | 0.41 / 0.40 | 0.73 / 0.91 | 0.53 / 0.33 | 0.74 (77) | 0.16 / 0.12 / −0.01 (67) |
| Any burden (seven burden constructs) | 108 | 0.40 (0.24 to 0.56) | 0.44 | 0.43 / 0.55 | 0.61 / 0.56 | 0.41 / 0.48 | 0.78 (66) | 0.27 / 0.36 / 0.52 (67) |
| *B. Patient-voice status, design-weighted to the full-text annotation (%, 95% CI)* |  |  |  |  |  |  |  |  |
| **Unit type** | **Units sampled: instrument-confirmed / not confirmed** | **Instrument-confirmed units confirmed by physician A** | **… by physician B** | **Unconfirmed units confirmed by physician A, n/N (Wilson 95% CI, unweighted)** | **… by physician B** | **Share of physician-confirmed voice captured by the instrument (A)** | **… (B)** |  |
| Forum articles | 37 / 15 | 75.7 (59.5–89.2) | 75.7 (62.2–89.2) | 9/15 (36–80) | 7/15 (25–70) | 51.8 (41.8–65.9) | 58.0 (45.9–74.9) |  |
| Push comments | 17 / 10 | 41.2 (17.6–64.7) | 58.8 (35.3–82.4) | 1/10 (2–40) | 0/10 (0–28) | 50.3 (20.2–100.0) | 100.0 (not estimable) |  |
| Ask-a-physician submissions | 41 / 5 | 98.9 (96.8–100.0) | 96.8 (93.6–100.0) | 4/5 (38–96) | 5/5 (57–100) | 97.6 (97.5–97.6) | 96.7 (96.6–96.8) |  |
| *C. Construct rates on the random sample of instrument-confirmed units (each source’s own confirmed units; design-weighted), and with the units the instrument missed* |  |  |  |  |  |  |  |  |
| **Construct** | **Instrument : articles / services, %** | **Physician A** | **Physician B** | **Services minus articles, pp (95% CI): instrument** | **Physician A** | **Physician B** | **Including missed units, physician A: articles / services, %** | **Physician B** |
| Information gap | 59.5 / 100.0 | 92.9 / 90.7 | 85.7 / 83.2 | +40.5 (24.3 to 56.8) | −2.2 (−15.7 to 11.1) | −2.5 (−20.2 to 15.3) | 96.3 / 89.2 | 91.7 / 82.0 |
| Emotional burden | 37.8 / 27.6 | 50.0 / 11.4 | 42.9 / 19.2 | −10.2 (−30.8 to 12.0) | −38.6 (−58.3 to −16.5) | −23.6 (−45.6 to −1.0) | 58.0 / 11.1 | 48.9 / 18.6 |
| **Construct** | **Instrument : articles / services, %** | **Physician A** | **Physician B** | **Services minus articles, pp (95% CI): instrument** | **Physician A** | **Physician B** | **Including missed units, physician A: articles / services, %** | **Physician B** |
| Medication experience | 27.0 / 16.8 | 39.3 / 26.2 | 14.3 / 12.6 | −10.3 (−28.4 to 9.3) | −13.0 (−36.9 to 9.7) | −1.7 (−18.7 to 14.8) | 36.4 / 25.6 | 14.3 / 12.2 |
| Lifestyle burden | 24.3 / 11.3 | 60.7 / 27.5 | 17.9 / 0.0 | −13.0 (−29.3 to 3.5) | −33.3 (−54.6 to −9.6) | −17.9 (−33.3 to −4.2) | 52.9 / 26.8 | 10.4 / 0.0 |
| System navigation | 10.8 / 5.1 | 28.6 / 16.9 | 14.3 / 5.3 | −5.7 (−18.2 to 5.4) | −11.6 (−34.0 to 8.3) | −9.0 (−24.0 to 4.8) | 25.5 / 16.5 | 14.3 / 5.1 |
| Access barrier | 0.0 / 0.0 | 17.9 / 6.6 | 0.0 / 0.0 | none labelled | −11.3 (−27.9 to 5.5) | none labelled | 30.7 / 6.4 | 0.0 / 0.0 |
| Caregiver burden | 5.4 / 3.0 | 25.0 / 3.6 | 10.7 / 0.0 | −2.4 (−10.8 to 6.2) | −21.4 (−39.1 to −4.4) | −10.7 (−23.8 to 0.0) | 18.3 / 3.5 | 12.2 / 0.0 |
| Cost burden | 2.7 / 0.0 | 32.1 / 6.8 | 3.6 / 0.0 | −2.7 (−8.1 to 0.0) | −25.4 (−44.2 to −6.7) | −3.6 (−11.5 to 0.0) | 32.7 / 6.6 | 2.1 / 0.0 |
| Any burden (seven burden constructs) | 56.8 / 47.6 | 89.3 / 60.1 | 57.1 / 28.7 | −9.2 (−30.9 to 15.2) | −29.2 (−47.5 to −10.2) | −28.4 (−52.0 to −4.7) | 89.1 / 58.6 | 69.1 / 27.8 |
| Caregiver authorship | 13.5 / 18.4 | 28.6 / 7.7 | 17.9 / 14.0 | +4.9 (−12.4 to 21.9) | −20.9 (−40.4 to −1.4) | −3.9 (−23.1 to 15.1) | 20.2 / 7.5 | 16.4 / 13.5 |
| *D. Instrument-rejected units by the instrument’s reason for rejection, and how many each physician confirmed* |  |  |  |  |  |  |  |  |
| **Unit type** | **Instrument’s reason** | **Units** | **Confirmed by physician A** | **… by physician B** | **… by both** |  |  |  |
| Forum articles | judged not relevant | 12 | 6 | 4 | 3 |  |  |  |
| Forum articles | relevant but not patient or caregiver voice | 3 | 3 | 3 | 3 |  |  |  |
| Push comments | judged not relevant | 9 | 1 | 0 | 0 |  |  |  |
| Push comments | relevant but not patient or caregiver voice | 1 | 0 | 0 | 0 |  |  |  |
| Ask-a-physician submissions | judged not relevant | 2 | 2 | 2 | 2 |  |  |  |
| Ask-a-physician submissions | relevant but not patient or caregiver voice | 3 | 2 | 3 | 2 |  |  |  |

Units: 160 sampled from the full-text annotation with a fixed seed (strata: disagree 10, construct 25, random 95, unconfirmed 30); each physician rated every unit independently, blind to model labels and to the other physician; there was no adjudication. κ intervals resample units (2,000 resamples); AC1 = Gwet’s first-order agreement coefficient, which is robust to skewed prevalence. Panel B weights each unit type’s sampled units by that unit type’s number of confirmed (or unconfirmed) units in the full-text annotation; intervals resample within strata (2,000 resamples). Captured share = confirmed units the physician confirms ÷ (those + unconfirmed units the physician confirms), each weighted to the full-text annotation. Panel C: a source’s rate uses the units that source judged patient or caregiver voice; the instrument’s rates here are on the sampled units, not the full-text annotation. Forum articles each physician confirmed: captured (random sample) any burden 89.3% and 57.1% (n = 28, 28); missed (unconfirmed stratum) 88.9% and 85.7% (n = 9, 7); caregiver-authored 28.6% and 17.9% captured vs 11.1% and 14.3% missed. Allocation of sampled units by stratum and unit type: disagree: KingNet 1, PTT articles 5, push comments 1, Taiwan e-Hospital 2, tcm.tw 1; construct: KingNet 7, PTT articles 8, push comments 2, Taiwan e-Hospital 4, tcm.tw 4; random: KingNet 12, PTT articles 37, push comments 17, Taiwan e-Hospital 14, tcm.tw 15; unconfirmed: KingNet 2, PTT articles 15, push comments 10, Taiwan e-Hospital 1, tcm.tw 2. The read frame is all units read in full by the full-text instrument; estimates describe those units, by unit type. † Interval not computed (fewer than five positives for a source). — = κ undefined because a source did not vary (e.g., every randomly sampled unit is instrument-confirmed). ‘Not estimable’ marks percentile intervals of zero width from strata too small to resample. Physicians A and B are two physician authors; which author is A is not disclosed.

**Table S7.** Production keyword pipeline against the full-text model: missing output for a broad construct and over-count of a narrow one (reference stratum; model-confirmed units)

| Domain and unit type | Units | Pipeline output stored, n (%) | Keyword rate, % | Model reference rate, % (95% CI) | Comparison |
|---|---|---|---|---|---|
| **Information gap, as stored (missing output counted as absent)** |  |  |  |  |  |
| All model-confirmed units | 2,272 | 680 (29.9) | 19.7 | 83.2 (81.6–84.7) | 4.2× lower (3.9–4.6) |
| — forum articles | 382 | 37 (9.7) | 5.8 | 58.6 (53.6–63.5) | 10.2× lower (7.2–16.2) |
| — ask-a-physician services | 1,656 | 630 (38.0) | 25.7 | 99.7 (99.3–99.9) | 3.9× lower (3.6–4.2) |
| **Information gap, units with a stored output** |  |  |  |  |  |
| — forum articles | 37 | 37 (100) | 59.5 (43.5–73.7) | 51.4 (35.9–66.6) | Similar (1.2× higher) |
| — KingNet | 630 | 630 (100) | 67.5 (63.7–71.0) | 100.0 (99.4–100.0) | 1.5× lower |
| — Taiwan e-Hospital and tcm.tw | 957 | 0 (0) | — | — | No output computed |
| **NHI navigation (24-term lexicon applied to every unit)** |  |  |  |  |  |
| All model-confirmed units | 2,272 | 2,272 (100) | 4.2 | 1.5 (1.0–2.0) | 2.9× higher (2.2–4.0) |
| — forum articles | 382 | 382 (100) | 8.4 | 1.6 (0.7–3.4) | 5.3× higher (3.0–17.0) |
| — ask-a-physician services | 1,656 | 1,656 (100) | 3.7 | 1.5 (1.0–2.2) | 2.5× higher (1.8–3.7) |

Information-gap reference = full-text model label; NHI-navigation reference = strict single-unit model read; neither is an adjudicated human label. The pipeline’s information-gap output was computed only for units within its aspect-processing scope (on PTT, mostly recent posts: median publication year 2024 for forum articles with an output vs 2014 without); the production outputs stored missing values as absent. Fold differences: reference ÷ keyword (lower) or keyword ÷ reference (higher), bootstrap 95% CI (2,000 unit resamples) where shown. Wilson 95% CIs.

### Multimedia Appendix 2. Verbatim Prompts for the Large Language Model Reads

Prompts are reproduced exactly as sent. The batched full-text instrument (A, B) was used for the primary read (gpt-6.1-sol, OpenAI Codex command-line interface), the end-to-end replica (Claude Sonnet 5.5, Claude Code command-line interface) — both in documents of 25 units — and the instrument pilot (25, 100 and 300 units); the single-unit prompts (C–E) are those of the single-unit GPT-5.5 reference read against which the full-text instrument was compared. The prompt of the strict single-unit NHI-navigation read is not reproduced here. No post text appears in this appendix.

#### A. System prompt of the batched full-text instrument

You are a labelling engine for a research study. You read Traditional Chinese health-forum text and return only JSON lines as instructed. Never use tools.

### B. Batched full-text annotation prompt: relevance and voice (Rubric A) and the eight constructs (Rubric B), one document of units as a JSON array

You will receive several independent posts as a JSON array of objects {“id“:…, “text“:…}. Apply BOTH rubrics below to EVERY post, judging each post strictly on its own text; one post must never influence another.

RUBRIC A (relevance and voice):

You are auditing whether a public Taiwanese online text unit (Traditional Chinese; it may be a forum article, a short forum comment, or a question submitted to an ask-a-physician service) is genuine patient voice. Read the FULL text. Reply ONLY with a JSON object exactly like: {“health_related“:0,“voice“:“other“}

health_related = 1 if the text concerns a specific person’s health, symptoms, diagnosis, tests, treatment, medication, or care (including a question put to a physician); 0 if it is general chat, opinion, jokes, news, advertising, policy debate, or not about anyone’s health.

voice = who is speaking about whose health: ‘patient’ (the author writes about their own health, symptoms, diagnosis, tests, treatment or care), ‘caregiver’ (the author writes about the health of a family member or a person they care for), ‘hcp’ (a clinician or professional answering or advising), ‘media’ (news, promotional, institutional, or reposted article), ‘other’ (general chat, opinion, a second-hand anecdote about a stranger, or not about anyone’s health).

RUBRIC B (constructs):

You are labelling a public Taiwanese health-forum post (Traditional Chinese) for eight binary friction constructs. Read the FULL text. Label 1 only when the construct is genuinely present for the author (or, for caregiver_burden, that the author is a caregiver voicing burden); passing mentions of vocabulary do not count. Reply with ONLY a JSON object exactly like:

{“info_gap“:0,“cost_burden“:0,“access_barrier“:0,“system_nav“:0,“emotional“:0,“medication_exp“:0,“lifestyle_burden“:0, “caregiver_burden“:0}

Definitions: info_gap = seeks information/advice, asks a clinical question, or expresses diagnostic/treatment uncertainty. cost_burden = voices actual financial strain from care costs. access_barrier = concrete difficulty reaching care (appointments, referrals, distance, waiting). system_nav = actively working through institutional processes (insurance coverage, eligibility, paperwork, which hospital/specialty). emotional = illness-tied emotional distress (fear, despair, exhaustion). medication_exp = first-hand medication experience, efficacy doubt or side-effects. lifestyle_burden = burden from illness-driven restriction of diet, activity, work, or sleep. caregiver_burden = the author cares for a patient and voices that burden.

OUTPUT FORMAT: for each post, output exactly one line containing one JSON object with the post’s id copied verbatim and every field, like

{“id“:“<id>”,“health_related“:0,“voice“:“other”,“info_gap“:0,“cost_burden“:0,“access_barrier“:0,“system_nav“:0,“emotion al“:0,“medication_exp“:0,“lifestyle_burden“:0,“caregiver_burden“:0}

Output exactly one line per post, in the same order as the input, and nothing else (no prose, no code fences, no numbering).

The documents were JSON arrays of objects {“id”, “text“}; the model returned one JSON line per unit. Units whose line was missing or unparseable were re-asked in smaller follow-up documents until every unit had a parseable line; no unit remained unread.

#### C. Single-unit relevance confirmation read (GPT-5.5; the single-unit reference read)

*Model: GPT-5.5 (OpenRouter route* openai/gpt-5.5). The prompt was sent as the user message, followed by the unit text *truncated at* 6,000 characters; outputs were health_related (0/1) and voice (patient, caregiver, hcp, media or other).

You are auditing whether a public Taiwanese online text unit (Traditional Chinese; it may be a forum article, a short forum comment, or a question submitted to an ask-a-physician service) is genuine patient voice. Read the FULL text. Reply ONLY with a JSON object exactly like: {“health_related“:0,“voice“:“other“}

health_related = 1 if the text concerns a specific person’s health, symptoms, diagnosis, tests, treatment, medication, or care (including a question put to a physician); 0 if it is general chat, opinion, jokes, news, advertising, policy debate, or not about anyone’s health.

voice = who is speaking about whose health: ‘patient’ (the author writes about their own health, symptoms, diagnosis, tests, treatment or care), ‘caregiver’ (the author writes about the health of a family member or a person they care for), ‘hcp’ (a clinician or professional answering or advising), ‘media’ (news, promotional, institutional, or reposted article), ‘other’ (general chat, opinion, a second-hand anecdote about a stranger, or not about anyone’s health).

POST:

#### D. Single-unit eight-construct read (GPT-5.5; the single-unit reference read)

*Model: GPT-5.5 (OpenRouter route* openai/gpt-5.5). The prompt was sent as the user message, followed by the post text *truncated at* 6,000 characters.

You are labelling a public Taiwanese health-forum post (Traditional Chinese) for eight binary friction constructs. Read the FULL text. Label 1 only when the construct is genuinely present for the author (or, for caregiver_burden, that the author is a caregiver voicing burden); passing mentions of vocabulary do not count. Reply with ONLY a JSON object exactly like:

{“info_gap“:0,“cost_burden“:0,“access_barrier“:0,“system_nav“:0,“emotional“:0,“medication_exp“:0,“lifestyle_burden“:0, “caregiver_burden“:0}

Definitions: info_gap = seeks information/advice, asks a clinical question, or expresses diagnostic/treatment uncertainty. cost_burden = voices actual financial strain from care costs. access_barrier = concrete difficulty reaching care (appointments, referrals, distance, waiting). system_nav = actively working through institutional processes (insurance coverage, eligibility, paperwork, which hospital/specialty). emotional = illness-tied emotional distress (fear, despair, exhaustion). medication_exp = first-hand medication experience, efficacy doubt or side-effects. lifestyle_burden = burden from illness-driven restriction of diet, activity, work, or sleep. caregiver_burden = the author cares for a patient and voices that burden.

POST:

#### E. Single-construct information-gap read (GPT-5.5; construct with physician reference labels)

*Model: GPT-5.5 (OpenRouter route* openai/gpt-5.5). The prompt was sent as the system message and the post text, truncated at 4,000 characters, as the user message.

Decide whether ONE health social-media post contains an INFORMATION GAP. Output STRICT JSON {“info_gap“:0|1}. info_gap=1 if the post seeks medical information or advice, asks a clinical question, or expresses uncertainty about a symptom, test, diagnosis, or treatment; else 0. Posts may be in Traditional Chinese, English, French, Japanese, or Simplified Chinese; judge in the original language. Output ONLY the JSON.

#### F. The 24-term NHI lexicon of the production keyword classifier (version 2.4)

健保 (NHI), 全民健保 (National Health Insurance), 健保卡 (NHI card), 健保署 (NHI Administration), 健保局 (NHI Bureau), 給付 (reimbursement), 不給付 (not reimbursed), 自費 (self-pay), 自付 (out-of-pocket), 部分負擔 (co-payment), 掛號費 (registration fee), 差額 (balance billing), 事前審查 (prior review), 事審 (prior review, short form), 給付規定 (reimbursement rules), 給付條件 (reimbursement criteria), 重大傷病 (catastrophic illness), 藥價 (drug price), 藥費 (drug cost), 新藥 (new drug), 轉診 (referral), 候診 (waiting to be seen), 排隊 (queueing), 等床 (waiting for a bed). A unit was counted as NHI navigation by the keyword instrument when any term matched after jieba segmentation.

### Multimedia Appendix 3. STROBE and TRIPOD-LLM Checklists

STROBE is applied as the closest fit for a cross-sectional content analysis of text units; TRIPOD-LLM items are those applicable to a large language model used as a measurement instrument rather than as a prediction model.

#### STROBE checklist (cross-sectional studies)

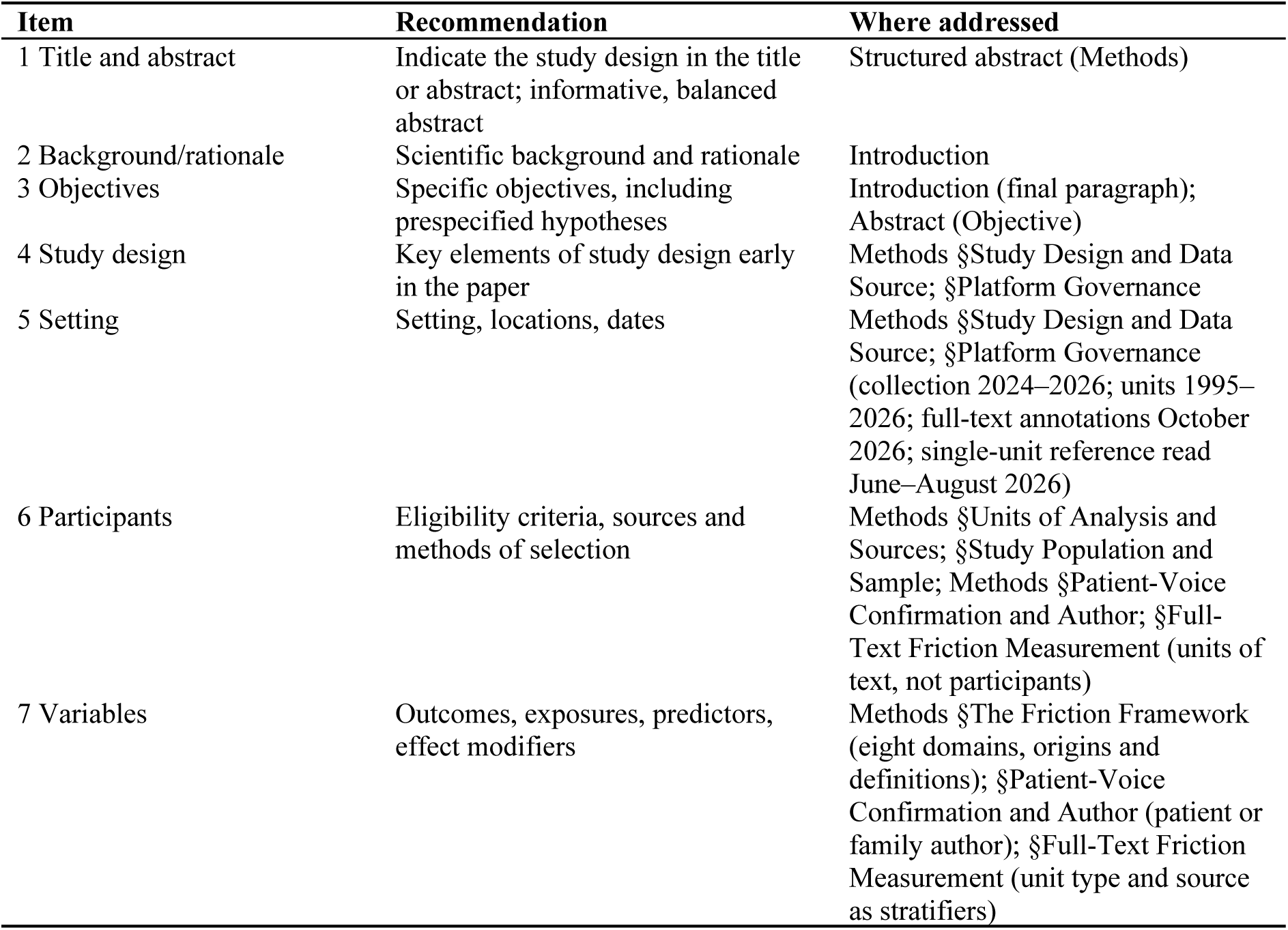
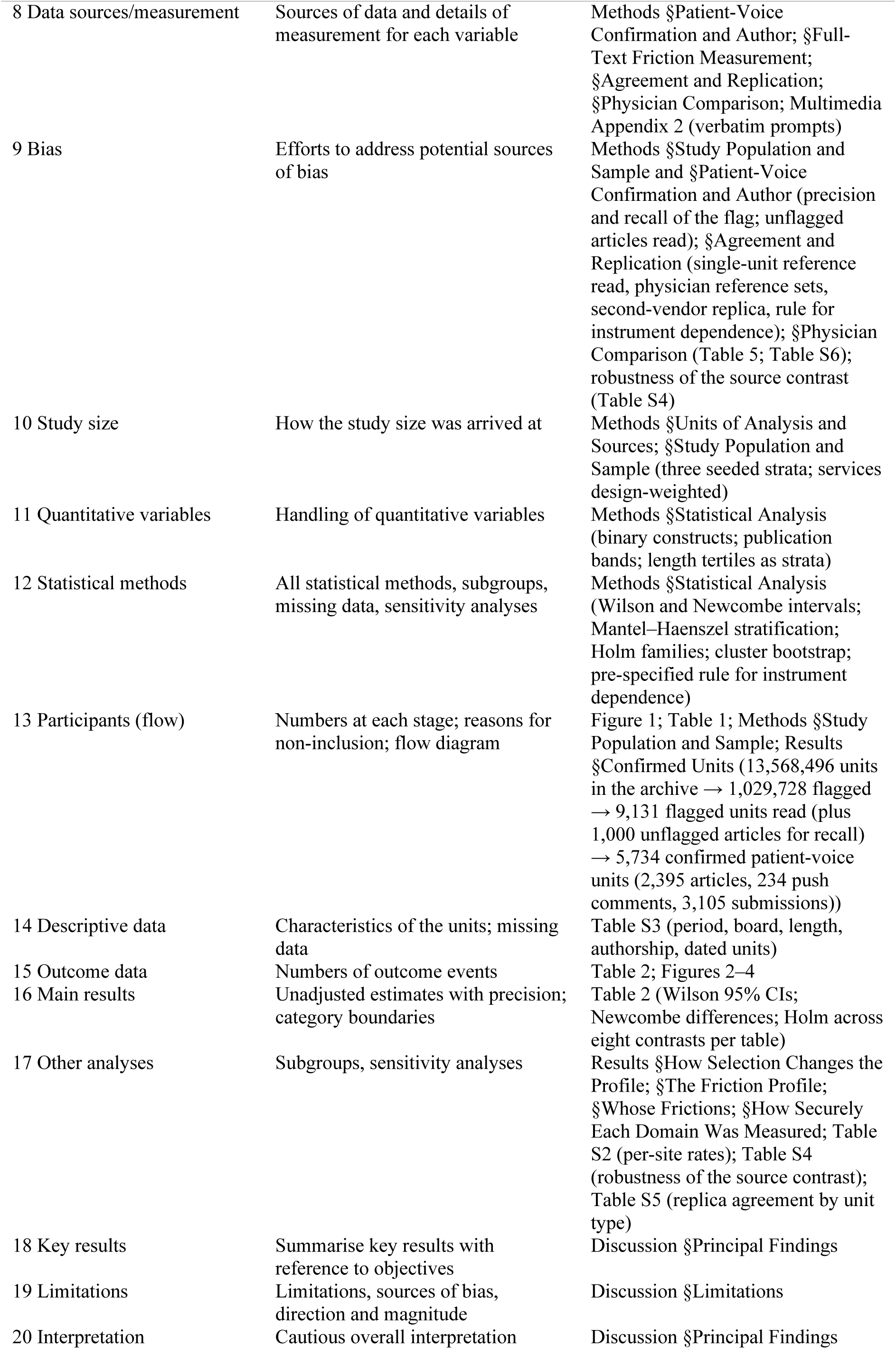
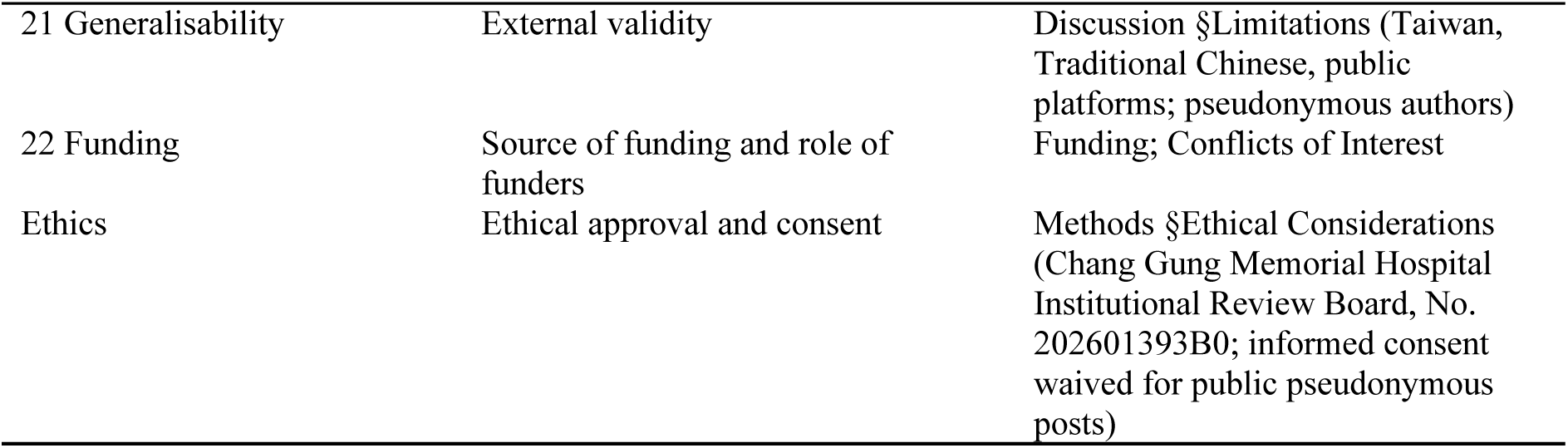

| Item | Recommendation | Where addressed |
|---|---|---|
| 1 Title and abstract | Indicate the study design in the title or abstract; informative, balanced abstract | Structured abstract (Methods) |
| 2 Background/rationale | Scientific background and rationale | Introduction |
| 3 Objectives | Specific objectives, including prespecified hypotheses | Introduction (final paragraph); Abstract (Objective) |
| 4 Study design | Key elements of study design early in the paper | Methods §Study Design and Data Source; §Platform Governance |
| 5 Setting | Setting, locations, dates | Methods §Study Design and Data Source; §Platform Governance (collection 2024–2026; units 1995–2026; full-text annotations October 2026; single-unit reference read June–August 2026) |
| 6 Participants | Eligibility criteria, sources and methods of selection | Methods §Units of Analysis and Sources; §Study Population and Sample; Methods §Patient-Voice Confirmation and Author; §Full-Text Friction Measurement (units of text, not participants) |
| 7 Variables | Outcomes, exposures, predictors, effect modifiers | Methods §The Friction Framework (eight domains, origins and definitions); §Patient-Voice Confirmation and Author (patient or family author); §Full-Text Friction Measurement (unit type and source as stratifiers) |
| 8 Data sources/measurement | Sources of data and details of measurement for each variable | Methods §Patient-Voice Confirmation and Author; §Full-Text Friction Measurement; §Agreement and Replication; §Physician Comparison; Multimedia Appendix 2 (verbatim prompts) |
| 9 Bias | Efforts to address potential sources of bias | Methods §Study Population and Sample and §Patient-Voice Confirmation and Author (precision and recall of the flag; unflagged articles read); §Agreement and Replication (single-unit reference read, physician reference sets, second-vendor replica, rule for instrument dependence); §Physician Comparison (Table 5; Table S6); robustness of the source contrast (Table S4) |
| 10 Study size | How the study size was arrived at | Methods §Units of Analysis and Sources; §Study Population and Sample (three seeded strata; services design-weighted) |
| 11 Quantitative variables | Handling of quantitative variables | Methods §Statistical Analysis (binary constructs; publication bands; length tertiles as strata) |
| 12 Statistical methods | All statistical methods, subgroups, missing data, sensitivity analyses | Methods §Statistical Analysis (Wilson and Newcombe intervals; Mantel–Haenszel stratification; Holm families; cluster bootstrap; pre-specified rule for instrument dependence) |
| 13 Participants (flow) | Numbers at each stage; reasons for non-inclusion; flow diagram | Figure 1; Table 1; Methods §Study Population and Sample; Results §Confirmed Units (13,568,496 units in the archive → 1,029,728 flagged → 9,131 flagged units read (plus 1,000 unflagged articles for recall) → 5,734 confirmed patient-voice units (2,395 articles, 234 push comments, 3,105 submissions)) |
| 14 Descriptive data | Characteristics of the units; missing data | Table S3 (period, board, length, authorship, dated units) |
| 15 Outcome data | Numbers of outcome events | Table 2; Figures 2–4 |
| 16 Main results | Unadjusted estimates with precision; category boundaries | Table 2 (Wilson 95% CIs; Newcombe differences; Holm across eight contrasts per table) |
| 17 Other analyses | Subgroups, sensitivity analyses | Results §How Selection Changes the Profile; §The Friction Profile; §Whose Frictions; §How Securely Each Domain Was Measured; Table S2 (per-site rates); Table S4 (robustness of the source contrast); Table S5 (replica agreement by unit type) |
| 18 Key results | Summarise key results with reference to objectives | Discussion §Principal Findings |
| 19 Limitations | Limitations, sources of bias, direction and magnitude | Discussion §Limitations |
| 20 Interpretation | Cautious overall interpretation | Discussion §Principal Findings |
| 21 Generalisability | External validity | Discussion §Limitations (Taiwan, Traditional Chinese, public platforms; pseudonymous authors) |
| 22 Funding | Source of funding and role of funders | Funding; Conflicts of Interest |
| Ethics | Ethical approval and consent | Methods §Ethical Considerations (Chang Gung Memorial Hospital Institutional Review Board, No. 202601393B0; informed consent waived for public pseudonymous posts) |

#### TRIPOD-LLM checklist (LLM as a text-labelling instrument)

| Item | Requirement | Where addressed |
|---|---|---|
| Title/abstract | Identify the use of an LLM and the task | Title; Abstract (Methods) |
| Background | Context and rationale for an LLM | Introduction |
| Objectives | Specify the LLM task | Abstract (Objective); Methods (relevance and voice classification; binary construct labelling) |
| Data source and eligibility | Describe the text corpus, dates and inclusion criteria | Methods §Study Design and Data Source; §Platform Governance; Methods §Units of Analysis and Sources; §Study Population and Sample |
| Reference standard | Define the reference standard and its provenance | Methods §Agreement and Replication; §Physician Comparison (single-unit GPT-5.5 reference read on the reference stratum; physician labels for information gap and NHI navigation); two physicians' independent labels of relevance, voice and all eight constructs on confirmed and unconfirmed units (Table 5; Table S6) |
| Model identification | Name, version, provider, access route, dates | Methods §Full-Text Friction Measurement (gpt-6.1-sol via the OpenAI Codex command-line interface, 23 October 2026; pilot of four models); §Agreement and Replication (Claude Sonnet 5.5 replica; GPT-5.5 single-unit reference read via OpenRouter, June–August 2026) |
| Prompting | Report prompts verbatim; system/user structure | Multimedia Appendix 2 (verbatim prompts) |
| Inference settings | Temperature, seed, token limits, truncation, batching | Methods §Full-Text Friction Measurement; §Agreement and Replication (provider defaults; no seed; 25 units per document for the primary read and the replica; seeded mixture of units within documents) |
| Output handling | Parsing, retries, failure rate | Methods §Full-Text Friction Measurement (one JSON object per unit; documents re-read until complete; every unit labelled) |
| Evaluation | Agreement metrics with CIs and denominators; stratified reporting | Results §How Securely Each Domain Was Measured; Tables 4 and 5; Tables S5 and S6 |
| Robustness | Prompt sensitivity, cross-model agreement, sensitivity analyses | Table 4 (second-vendor rates and κ by domain); Table S5 (agreement); rule for instrument dependence; Table S4 |
| Fairness and subgroup performance | Performance across relevant subgroups | Results §How Securely Each Domain Was Measured (agreement by unit type / platform); no demographic subgroups exist for pseudonymous posts |
| Data leakage / training contamination | Possibility that evaluation text was in training data | Discussion §Limitations (public posts 1995–2026 may be present in training data) |
| Human oversight | Role of humans in labelling and interpretation | Methods §Agreement and Replication; §Physician Comparison (physician reference sets); Limitations |
| Costs and environmental impact | Report where possible | Primary read and replica through command-line interfaces; single-unit reference reads through the OpenRouter application programming interface; costs and compute not reported |
| Availability | Code, prompts, outputs, model access | Data Availability (on reasonable request); Multimedia Appendix 2 (verbatim prompts) |
| Generative AI in writing | Disclose use in manuscript preparation | Generative AI Disclosure |

## References

1. Eysenbach G. Infodemiology and infoveillance: framework for an emerging set of public health informatics methods to analyze search, communication and publication behavior on the internet. J Med Internet Res. 2009;11(1):e11. doi:10.2196/jmir.1157

2. Mavragani A. Infodemiology and infoveillance: scoping review. J Med Internet Res. 2020;22(4):e16206. doi:10.2196/16206

3. Sarker A, Ginn R, Nikfarjam A, et al. Utilizing social media data for pharmacovigilance: a review. J Biomed Inform. 2015;54:202–212. doi:10.1016/j.jbi.2015.02.004

4. Schmidt AL, Rodriguez-Esteban R, Gottowik J, Leddin M. Applications of quantitative social media listening to patient-centric drug development. Drug Discov Today. 2022;27(5):1523–1530. doi:10.1016/j.drudis.2022.01.015

5. Cimiano P, Collins B, De Vuono MC, et al. Patient listening on social media for patient-focused drug development: a synthesis of considerations from patients, industry and regulators. Front Med (Lausanne). 2024;11:1274688. doi:10.3389/fmed.2024.1274688

6. Cimiano P, Brazda N, Hartung M, et al. Best practices for the collection and analysis of patient experience data from social media for patient-focused drug development. Front Med (Lausanne). 2026;12:1703923. doi:10.3389/fmed.2025.1703923

7. US Food and Drug Administration. Patient-focused drug development: collecting comprehensive and representative input. Guidance for industry, Food and Drug Administration staff, and other stakeholders. Silver Spring, MD: US Food and Drug Administration; June 2020. https://www.fda.gov/regulatory-information/search-fda-guidance-documents/patient-focused-drug-development-collecting-comprehensive-and-representative-input. Accessed 19 October 2026.

8. Chiu MP. An investigation of the questions posted on medical consultation websites. Health Info Libr J. 2016;33(4):283–294. doi:10.1111/hir.12153

9. Hong Z, Deng Z, Evans R, Wu H. Patient questions and physician responses in a Chinese health Q&A website: content analysis. J Med Internet Res. 2020;22(4):e13071. doi:10.2196/13071

10. Wang RH, Hong YW, Li CC, et al. Using social media data in diabetes care: bridging the conceptual gap between health providers and the network population. BMC Prim Care. 2022;23(1):241. doi:10.1186/s12875-022-01846-0

11. Jeon H, Yu SY, Chertkova O, et al. Real-world insights of patient voices with age-related macular degeneration in the Republic of Korea and Taiwan: an AI-based Digital Listening study by Semantic-Natural Language Processing. BMC Med Inform Decis Mak. 2025;25(1):137. doi:10.1186/s12911-025-02929-5

12. Hao H, Zhang K. The voice of Chinese health consumers: a text mining approach to web-based physician reviews. J Med Internet Res. 2016;18(5):e108. doi:10.2196/jmir.4430

13. Li J, Yang Y, Mao C, et al. Revealing patient dissatisfaction with health care resource allocation in multiple dimensions using large language models and the International Classification of Diseases 11th Revision: aspect-based sentiment analysis. J Med Internet Res. 2025;27:e66344. doi:10.2196/66344

14. Li J, Yang Y, Chen R, et al. Identifying healthcare needs with patient experience reviews using ChatGPT. PLoS One. 2025;20(3):e0313442. doi:10.1371/journal.pone.0313442

15. Mao C, Li J, Pang PCI, Zhu Q, Chen R. Identifying kidney stone risk factors through patient experiences with a large language model: text analysis and empirical study. J Med Internet Res. 2025;27:e66365. doi:10.2196/66365

16. Cheng TM. Taiwan’s new national health insurance program: genesis and experience so far. Health Aff (Millwood). 2003;22(3):61–76. doi:10.1377/hlthaff.22.3.61

17. Shih Y, Liao K, Chen Y, Lin F, Hsiao F. Reimbursement lag of new drugs under Taiwan’s National Health Insurance system compared with United Kingdom, Canada, Australia, Japan, and South Korea. Clin Transl Sci. 2020;13(5):916–922. doi:10.1111/cts.12778

18. Yin JDC. Vaccine hesitancy in Taiwan: temporal, multilayer network study of echo chambers shaped by influential users. Online J Public Health Inform. 2024;16:e55104. doi:10.2196/55104

19. Lazer D, Kennedy R, King G, Vespignani A. The parable of Google Flu: traps in big data analysis. Science. 2014;343(6176):1203–1205. doi:10.1126/science.1248506

20. Lamb A, Paul MJ, Dredze M. Separating fact from fear: tracking flu infections on Twitter. In: Proceedings of the 2013 Conference of the North American Chapter of the Association for Computational Linguistics: Human Language Technologies. Atlanta, GA: Association for Computational Linguistics; 2013:789–795. https://aclanthology.org/N13-1097/. Accessed 20 August 2026.

21. Nikfarjam A, Sarker A, O’Connor K, Ginn R, Gonzalez G. Pharmacovigilance from social media: mining adverse drug reaction mentions using sequence labeling with word embedding cluster features. J Am Med Inform Assoc. 2015;22(3):671–681. doi:10.1093/jamia/ocu041

22. Tufekci Z. Big questions for social media big data: representativeness, validity and other methodological pitfalls. Proc Int AAAI Conf Web Soc Media. 2014;8(1):505–514. doi:10.1609/icwsm.v8i1.14517

23. D’Souza RS, Hooten WM, Murad MH. A proposed approach for conducting studies that use data from social media platforms. Mayo Clin Proc. 2021;96(8):2218–2229. doi:10.1016/j.mayocp.2021.02.010

24. Gilardi F, Alizadeh M, Kubli M. ChatGPT outperforms crowd workers for text-annotation tasks. Proc Natl Acad Sci U S A. 2023;120(30):e2305016120. doi:10.1073/pnas.2305016120

25. Deiner MS, Honcharov V, Li J, Mackey TK, Porco TC, Sarkar U. Large language models can enable inductive thematic analysis of a social media corpus in a single prompt: human validation study. JMIR Infodemiology. 2024;4:e59641. doi:10.2196/59641

26. Hairston J, Ranjan R, Lakamana S, et al. Automating inductive thematic analyses of health content using large language models: a proof-of-concept study using social media data. JAMIA Open. 2025;8(5):ooaf102. doi:10.1093/jamiaopen/ooaf102

27. Zhou X, Zhou J, Wang C, et al. A suite of large language models for public health infoveillance. NPJ Digit Med. 2026;9(1):270. doi:10.1038/s41746-026-02435-6

28. Guo Y, Ovadje A, Al-Garadi MA, Sarker A. Evaluating large language models for health-related text classification tasks with public social media data. J Am Med Inform Assoc. 2024;31(10):2181–2189. doi:10.1093/jamia/ocae210

29. Pangakis N, Wolken S, Fasching N. Automated annotation with generative AI requires validation. arXiv. Preprint posted online May 31, 2023. doi:10.48550/arXiv.2306.00176

30. Törnberg P. Best practices for text annotation with large language models. arXiv. Preprint posted online February 5, 2024. doi:10.48550/arXiv.2402.05129

31. Barrie C, Palaiologou E, Törnberg P. Prompt stability scoring for text annotation with large language models. arXiv. Preprint posted online July 2, 2024. doi:10.48550/arXiv.2407.02039

32. Sanders R, Linn AJ, Araujo TB, Vliegenthart R, van Eenbergen MC, van Weert JCM. Different platforms for different patients’ needs: automatic content analysis of different online health information platforms. Int J Hum Comput Stud. 2020;137:102386. doi:10.1016/j.ijhcs.2019.102386

33. Le LH, Hoang PA, Pham HC. Sharing health information across online platforms: a systematic review. Health Commun. 2023;38(8):1550–1562. doi:10.1080/10410236.2021.2019920

34. Roberts K, Demner-Fushman D. Interactive use of online health resources: a comparison of consumer and professional questions. J Am Med Inform Assoc. 2016;23(4):802–811. doi:10.1093/jamia/ocw024

35. Kessler SH, Schmidt-Weitmann S. Diseases and emotions: an automated content analysis of health narratives in inquiries to an online health consultation service. Health Commun. 2021;36(2):226–235. doi:10.1080/10410236.2019.1673950

36. von Elm E, Altman DG, Egger M, Pocock SJ, Gøtzsche PC, Vandenbroucke JP. The Strengthening the Reporting of Observational Studies in Epidemiology (STROBE) statement: guidelines for reporting observational studies. Lancet. 2007;370(9596):1453–1457. doi:10.1016/S0140-6736(07)61602-X

37. Gallifant J, Afshar M, Ameen S, et al. The TRIPOD-LLM reporting guideline for studies using large language models. Nat Med. 2025;31(1):60–69. doi:10.1038/s41591-024-03425-5

38. Gwet KL. Computing inter-rater reliability and its variance in the presence of high agreement. Br J Math Stat Psychol. 2008;61(Pt 1):29–48. doi:10.1348/000711006X126600

39. Wilson EB. Probable inference, the law of succession, and statistical inference. J Am Stat Assoc. 1927;22(158):209–212. doi:10.1080/01621459.1927.10502953

40. Newcombe RG. Interval estimation for the difference between independent proportions: comparison of eleven methods. Stat Med. 1998;17(8):873–890. doi:10.1002/(SICI)1097-0258(19980430)17:8<873::AID-SIM779>3.0.CO;2-I

41. Holm S. A simple sequentially rejective multiple test procedure. Scand J Stat. 1979;6(2):65–70.

42. Cohen J. A coefficient of agreement for nominal scales. Educ Psychol Meas. 1960;20(1):37–46. doi:10.1177/001316446002000104

43. Feinstein AR, Cicchetti DV. High agreement but low kappa: I. The problems of two paradoxes. J Clin Epidemiol. 1990;43(6):543–549. doi:10.1016/0895-4356(90)90158-L

44. Cicchetti DV, Feinstein AR. High agreement but low kappa: II. Resolving the paradoxes. J Clin Epidemiol. 1990;43(6):551–558. doi:10.1016/0895-4356(90)90159-M

45. Eysenbach G, Till JE. Ethical issues in qualitative research on internet communities. BMJ. 2001;323(7321):1103–1105. doi:10.1136/bmj.323.7321.1103

46. Conway M. Ethical issues in using Twitter for public health surveillance and research: developing a taxonomy of ethical concepts from the research literature. J Med Internet Res. 2014;16(12):e290. doi:10.2196/jmir.3617

